# Paired Airway and Blood Multiomics Defines Coordinated Immune Adaptation Across the Airway–Blood Axis in Preterm Infants

**DOI:** 10.64898/2026.08.17.26360603

**Authors:** Safwen Kadri, Zhenda Wang, Claudia Nussbaum, Johannes B. Müller-Reif, Alicia-Sophie Schebesta, Rizqah Kamies, Brittany T. Rupp, Tanja Seegmüller, Caroline Johansson, Maarten Weiß, Axel Heep, Eduard Malik, Kai Förster, Andreas Flemmer, Mapping the Inhalation Interface Network (MPII-NET), Karin Loser, Herbert B. Schiller, Kevin Matthew Byrd, Anne Hilgendorff

## Abstract

Immune adaptation after birth requires coordinated remodeling across the airway– blood axis, yet how these compartments communicate during early postnatal life remains poorly understood. In preterms, dysregulation of this immune response determines mortality and morbidity. We performed paired single-cell RNA sequencing (scRNA-seq) and mass spectrometry-based proteomic profiling of airway samples (deep pharyngeal aspirates, DPA) and matched whole blood from 19 neonates spanning extreme preterm (<28 weeks) to term gestation, sampled at two postnatal timepoints (1–3 days and 4–10 days). This integrated multiomic atlas revealed coordinated and compartment-specific immune adaptation across the airway–blood axis during the first week of life. We observed gestational age-dependent shifts in cell composition in both compartments, including expansion of immature hematopoietic and myeloid populations in blood and distinct myeloid and epithelial programs in DPA, accompanied by compartment-specific inflammatory and innate immune gene expression that evolved during the first week of life. Unexpectedly, we identified a circulating respiratory epithelial-like cell population in neonatal blood whose abundance correlated with prematurity and lung disease and which we validated by flow cytometry as well as in independent datasets. Matched plasma proteomics revealed a gestational age axis and a disease-associated axis; an Organ-to-System Score derived from lung-restricted plasma proteins tracked lung injury severity and distinguished trajectories toward chronic lung disease as early as 72 hours after birth. Together, these multiomic data describe coordinated and divergent immune programs across mucosal and systemic compartments in early preterm life, identify circulating respiratory epithelial-like cells as a candidate blood-accessible signal of airway–blood interface perturbation, and prioritize ciliated–myeloid signaling (LAMA5–ITGB1) as a candidate axis underlying neonatal lung disease. This systems-level framework provides a platform for biomarker discovery and mechanistic studies in larger cohorts.

## Introduction

At birth, the immune system transitions from a comparatively protected intrauterine environment to continuous postnatal exposure to oxygen, microbes, nutrients, and the mechanical forces of the first breath and circulatory transition. Term neonates navigate this transition with maternal support and a physiologically prepared immune-respiratory system^1^. Upon premature birth - a leading driver of neonatal mortality and long-term morbidity worldwide^2,3^- the coordinated adaptation across the airway–blood axis that is required to successfully conquer the transition from fetal to postnatal life, faces the challenge of functional and structural immaturity. Preterm infants face altered immune responses that increase the risk of infection, sustained inflammation, and the development of long-term complications such as chronic lung disease^3,4^. The first week of life represents a critical developmental window during which early immune and respiratory adaptation strongly influence subsequent morbidity and long-term health ^5–7^. Despite the clinical significance, the molecular architecture of early-life immune adaptation and the coordination of immune programs across body compartments remains incompletely understood.

Cardiopulmonary adaptation is central to postnatal transition. In preterm infants, early mortality and morbidity are strongly shaped by the ability of the immature respiratory system to adapt to extrauterine life, and respiratory outcomes remain among the strongest predictors of long-term health^2,3^. Insults such as oxygen exposure, invasive and non-invasive ventilation, infections and microbial colonization, and the subsequent development of sustained inflammation and tissue injury each has the potential to reshape coordinated local and systemic immune responses. Neonatal respiratory distress syndrome (RDS) and bronchopulmonary dysplasia (BPD) reflect dysregulated responses of the immature lung and immune system to these early postnatal challenges^4,8–10^.

Whether airway and systemic immune compartments mature as coordinated or independent systems during early postnatal life remains poorly understood. Most prior studies have profiled either circulating blood or airway-derived samples in isolation, rarely with simultaneous single-cell and proteomic measurements at multiple postnatal timepoints^1,11–14^. As a consequence, it remains unclear whether immune changes in the airway and blood reflect compartment-specific responses or a larger mucosal– systemic program, which - when disturbed - primes the host immune response beyond the individual organ or distinct event. Defining these coordinated immune trajectories is essential for interpreting neonatal biomarkers, understanding disease pathogenesis, and developing compartment-aware therapeutic strategies.

Here, we profile immune adaptation across the first week of life in 19 neonates spanning extreme preterm to term gestation. We integrated single-cell RNA sequencing and proteomic profiling of paired deep pharyngeal aspirates (DPA) and blood at two early postnatal timepoints, enabling simultaneous analysis of gestational age-dependent immune signatures, early postnatal remodeling, and compartment-specific versus shared programs. Matched plasma and DPA proteomics linked transcriptional cell states to circulating immune and tissue-remodeling signals.

We hypothesized that early-life immune adaptation is a coordinated process across airway and blood compartments that is shaped by both maturity and subsequent perturbations including infection and lung disease. Using this framework, we define coordinated and compartment-specific immune trajectories across the airway–blood axis, identify candidate mechanisms linking epithelial and myeloid remodeling to neonatal lung disease, and establish a multiomic framework for understanding early-life immune adaptation. Confirmation of key findings in independent datasets and a plasma proteomic signature capturing the outlined biology in a blood-accessible framework that may predict respiratory outcomes strengthen the study and its clinical implication.

## Methods

### Patient recruitment and sampling

We prospectively enrolled 19 neonates with gestational ages spanning extreme preterm to term (from 23.9 to 41.6 weeks gestational age) recruited between 06/2023 and 06/2024 at the Perinatal Center of the Ludwig-Maximilian University, as part of the AIRR (Attention to Infants at Respiratory Risks) study (LMU Ethics Board #195-07; German Registry for Clinical Studies DRKS00004600). Serial samples were collected during the first 3 days of life and between days of life 4 and 10 (**Table 1**). Infants with severe congenital malformations, chromosomal abnormalities, inborn errors of metabolism, or a decision for palliative therapy immediately after birth were excluded, consistent with the inclusion/exclusion criteria previously published^15^. Clinical and laboratory variables were collected from birth to discharge (**Table 1**).

**Table 1:**
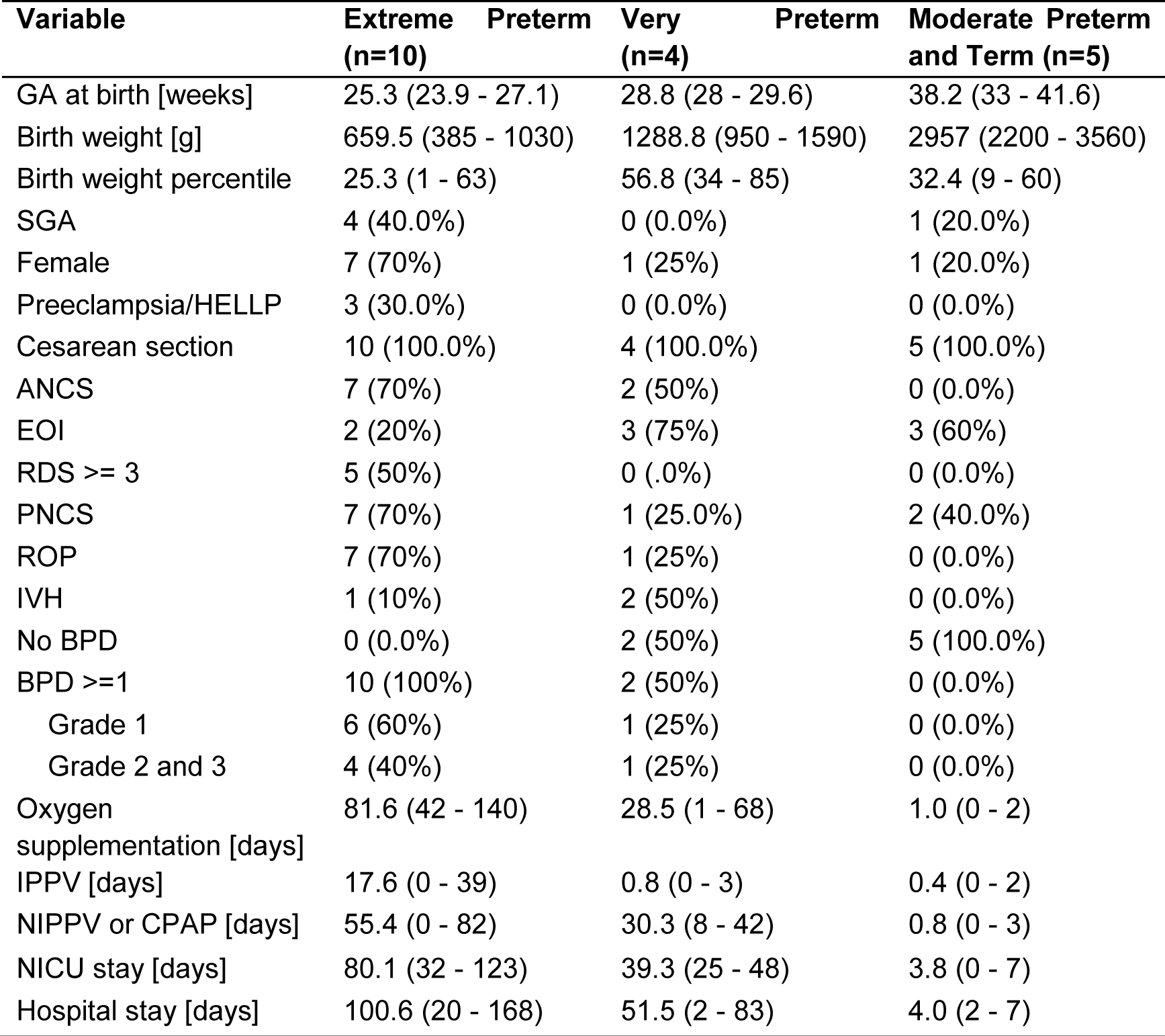
Cohort Characteristics. ANCS, Antenatal corticosteroids; BPD^2^, Bronchopulmonary Dysplasia; EOI, Early onset infection; GA, Gestational Age; IPPV, Invasive Positive Pressure Ventilation; HELPP, Hemolysis, Elevated Liver enzymes, and Low Platelets; IVH^68^, Intraventricular Hemorrhage; NIPPV, Non-invasive Positive Pressure Ventilation; PNCS, Postnatal corticosteroids; RDS, Respiratory Distress Syndrome (not available in n = 1 infant in mild BPD group); ROP, Retinopathy of prematurity (not available in n = 1 infant in no, mild and moderate/severe group, respectively); SGA, Small for gestational age, i.e. birth weight below 10th percentile; one patient transferred.

Comprehensive clinical monitoring was performed until discharge from neonatal care including data obtained from the respective pregnancies. The classification of immaturity followed the criteria established by the World Health Organization^16,17^. Age is presented in weeks gestation at birth. Being born small for gestational age (SGA) was defined as birth weight <10th percentile. Chorioamnionitis was defined as inflammatory alterations of the chorionic plate (histology) or clinical and laboratory signs of maternal and fetal inflammation/infection^18^. Postnatally, systemic infections were diagnosed based on one or more clinical and laboratory signs of infection^19^ and classified as early onset infections when the onset was within the first 72 hours of life. Postnatally, diagnosis and severity of respiratory distress syndrome (RDS) were scored on anterior-posterior chest radiographs according to Couchard et al.^20^. ROP, retinopathy of prematurity. IVH, intraventricular hemorrhage. PDA, patent ductus arteriosus; PH indication, signs of pulmonary hypertension. IPPV, invasive positive pressure ventilation; NIPPV, non-invasive positive pressure ventilation; O2 supplementation, oxygen supplementation; PNCS, postnatal corticosteroids; ICU, intensive care unit. BPD severity followed the criteria established by the National Institute of Child Health and Human Development (NICHD) workshop, categorizing infants into one of three groups: mild BPD was characterized by the requirement for oxygen (O2) supplementation for 28 days, with no need for O2 supplementation at 36 weeks postmenstrual age (PMA); moderate BPD entailed O2 supplementation with a fraction of inspired oxygen (FiO2) below 0.30 at 36 weeks PMA; severe BPD was diagnosed when infants required O2 supplementation with FiO2 exceeding 0.30 at 36 weeks PMA and/or positive pressure ventilation/continuous positive pressure^21^.

Blood samples and deep pharyngeal aspirates (DPA) were obtained during routine procedures in the first half of the day. Peripheral venous blood samples were obtained according to clinical standard and harvested into neonatal EDTA tubes (Sarstedt, Germany); DPA were acquired during routine suctioning of the airway at the end of the soft palate and harvested into a sterile Eppendorf tube (Eppendorf, Germany) (**Supplementary Table S1**).

Confirmatory flow cytometry data were acquired from umbilical cord blood samples from n=59 neonates (n=31 preterm (from 25.3 to 35.0 weeks gestational age) and 28 term (from 37.1 to 39.3 weeks gestational age), at the Perinatal Center, Oldenburg Hospital (UOL Ethics Board #2021-116, German Registry for Clinical Studies DRKS00025864; UOL Ethics Board #2024-124, German Registry for Clinical Studies DRKS00036823) following the in- and exclusion criteria outlined above.

### Sample processing

For each neonate, we aimed at collecting matched peripheral blood and DPA samples at two distinct postnatal time points (timepoint 1 (day of life 1-3) and timepoint 2 (day of life 4-10)). In some cases, clinical constraints led to sample attrition resulting in complete and partially complete longitudinally paired sample sets (**Supplementary Table S1**).

Peripheral venous blood samples were diluted with DPBS at 1:1 ratio, and gently applied overlaying onto 10ml Lymphoprep (STEMCELL, 07861) in a Protein LoBind 15ml conical tube (Eppendorf, 0030122216). Centrifuge at 450 x g for 20 minutes at room temperature, with minimal acceleration rate and brake off.

DPA samples were diluted with DPBS at 1:1 ratio, and mechanically dissociated by gently pipetting. The mix was filtered through a 70μm MACS strainer and collected with a Protein LoBind 15ml conical tube. Centrifuge at 450 x g for 10 minutes at room temperature.

The upper plasma layer and the upper supernatant layer were collected and frozen separately. The mononuclear cell layer and the pellet layer were retained, washed once with DPBS, counted with Neubauer hemocytometers and cryopreserved with 10% DMSO and 90% FBS.

For PBMC isolation and flow cytometry, umbilical cord blood samples were harvested in EDTA tubes (Sarstedt, Germany) in the delivery room before transfer into tubes with Tissue Storage Solution (Miltenyi-Biotec, 1:5 dilution) for sample preservation.

### Single cell analysis

#### Library preparation and sequencing

Cryopreserved cells were fixed with Evercode™ Cell Fixation v2 (Parse Biosciences) with user manual v2.1.1; Library was prepared with Evercode™ WT v2 (Parse Biosciences) with user manual v2.2.1. The quantified and quality controlled library was sequenced using NovaSeq 6000 System with S1 Reagent Kit v1.5 of 200 cycles (Illumina, 20028318) at Core Facility Genomics of Helmholtz Munich.

#### Read alignment and gene count matrix generation

Raw FASTQ files were aligned and gene-count matrices were generated using the Parse Biosciences split-pipe pipeline v1.0.6 with default parameters against the human reference genome (GRCh38).

#### Quality control and filtering

Per-cell quality metrics — total UMI counts (total_counts), number of genes detected (n_genes_by_counts), and percentage of mitochondrial (pct_counts_mt), ribosomal (pct_counts_ribo), and hemoglobin gene (pct_counts_hb) reads, were computed using scanpy (v1.11.4)^22^. Outlier cells were flagged using median-absolute-deviation (MAD)-based thresholds: cells were excluded if log10(total_counts) or log10(n_genes_by_counts) deviated by more than 5 MADs from the per-sample median, if pct_counts_mt exceeded 15%, or if fewer than 200 genes were detected. Doublets were identified using scDblFinder^23^ (v1.16.0) with per-sample scoring; per-cell scDblFinder scores were retained as a metadata covariate. Ambient RNA contamination was assessed and corrected using SoupX (v4.0.4). After quality filtering and doublet removal, a total of 49,733 cells were retained for downstream analysis.

#### Normalization, feature selection, and dimensionality reduction

Log-normalized counts (total counts scaled to 10,000 per cell followed by natural-log transformation; scanpy.pp.normalize_total and scanpy.pp.log1p) were used for all downstream analyses. Highly variable genes (HVGs) were selected using scanpy.pp.highly_variable_genes across the merged DPA and PBMC datasets. Principal component analysis (PCA) was performed on the HVG-scaled expression matrix (top 50 PCs). Batch effects across donors and compartments were corrected using Harmony^24^ (v0.1.8) applied to the PCA embedding. A k-nearest-neighbor (kNN) graph was constructed on the Harmony-corrected embedding (scanpy.pp.neighbors) and used for UMAP visualization (scanpy.tl.umap) and Leiden clustering (scanpy.tl.leiden).

#### Cell type annotation

Cell types were annotated iteratively by cross-referencing cluster-specific marker gene expression against canonical lineage markers for epithelial, myeloid, and lymphoid populations. Annotation was refined over successive Leiden clustering rounds until stable cluster identities were achieved, further guided by neonatal-specific cell type markers established in published neonatal lung single-cell datasets^11,25–28^ and cross-referenced to published single-cell data sets (GSE236099^11^). DPA and PBMC datasets were annotated independently and subsequently merged into a combined object for cross-compartment analyses, with compartment of origin (DPA vs. PBMC) retained as a metadata covariate throughout downstream analyses.

#### Pathway activity and module scoring

Gene expression module scores were computed using scanpy.tl.score_genes, which scores each cell relative to a background set of randomly sampled control genes matched for mean expression. Gene sets were drawn from Hallmark, Gene Ontology Biological Process (GOBP), and curated literature-based gene lists for biologically relevant programs including IFN response, TNF–NF-κB signaling, phagocytosis, antigen presentation, complement activation, chemokine signaling, M1/M2 polarization, glycolysis, oxidative phosphorylation, tissue remodeling, oxidative stress, mucociliary differentiation, ciliopathy/intraflagellar transport, epithelial barrier, and lung development programs.

Disease associations between per-cell module scores and clinical variables (GA, RDS, BPD, EOI) were assessed using linear mixed models (LMMs) fitted per cell type with donor as a random effect, using the lme4 package in R v4.5.0. β coefficients and nominal p-values were reported alongside Benjamini–Hochberg (BH) adjusted q-values within each test family.

#### Cell-type importance

To quantify the contribution of each annotated cell type to postnatal immune remodeling, a composite importance score was computed per cell type × gestational maturity group in DPA and PBMC independently. For each combination, two components were calculated between timepoint 1 (day of life 1-3) and timepoint 2 (day of life 4-10) : (i) a frequency change, defined as log₂(f_TP2 / f_TP1), where f_TP1 and f_TP2 are the fractions of that cell type among all cells of that maturity group at each timepoint; and (ii) an expression change, defined as the mean absolute log₂ fold-change across the top 500 most variable genes (ranked by cross-cell variance) between TP1 and TP2 cells of that type. The importance score was computed as the geometric mean of the absolute frequency change and the expression change: importance = √(|log₂FC_frequency| × mean|log₂FC_expression|). Scores were normalized within each compartment (PBMC and DPA) to the maximum observed importance score. Cell types with fewer than 10 cells at either timepoint were excluded. Results were visualized as a dot plot in which the x-axis encodes the direction and magnitude of frequency change (filled dots = expanding, hollow = contracting), dot size encodes normalized importance, and an inner white dot encodes the relative expression change magnitude.

#### Cell-type–metadata association radar plots

To visualize the relative contribution of clinical variables to transcriptional variation within each annotated cell type, a metadata association score was computed per cell type × clinical variable combination in PBMC and DPA independently. For each cell type and each metadata variable (gestational age, BPD severity, RDS severity, and EOI), the mean coefficient of determination (mean R²) was calculated across all genes by fitting per-gene linear regression models regressing log-normalized gene expression onto the metadata variable across all cells of that type. The resulting scores were normalized per cell type to yield a relative profile across the four clinical axes. Results were visualized as radar plots in which each axis represents one clinical variable and the area covered reflects the overall transcriptional association of that cell type with each clinical dimension. Cell types with fewer than 10 cells were excluded. Analyses were performed separately for PBMC and DPA compartments and at each postnatal timepoint.

#### Pseudobulk differential expression

Pseudobulk differential expression between conditions (gestational age groups, timepoints, disease strata) was performed by summing raw counts per donor per cell type using DESeq2^29^(v1.42.1), with donor as the unit of replication. Genes were considered differentially expressed at BH-adjusted p < 0.05.

#### Myeloid subclustering

Myeloid cells from integrated PBMC and DPA (n = 3,977) were isolated by subsetting on classical monocytes, non-classical monocytes, macrophages, plasmacytoid dendritic cells, neutrophils, and DPA monocyte/macrophage populations. The subset was reprocessed with HVG selection, PCA, Harmony batch correction, kNN graph construction, and Leiden clustering at increased resolution to resolve myeloid subpopulations. Subclusters were annotated using marker genes: IL1R2+ homeostatic classical monocytes, VCAN+ migratory classical monocytes, and CD16+ non-classical monocytes in PBMC; and Mono_Recruited, Mono_Inflammatory, Mono_AM-like, Mono_Stressed, Mono_Transit, and Mono_GranLike in DPA.

#### Myeloid trafficking analysis

Myeloid cell proportions were computed per donor as the fraction of myeloid cells relative to all cells at each timepoint. A trafficking score was defined per donor as DPA myeloid% − PBMC myeloid%, where positive values indicate lung enrichment relative to blood. Compartment-specific myeloid gene signatures were identified by Wilcoxon rank-sum differential expression (BH-adjusted p < 0.05) comparing myeloid versus all other cells independently in PBMC and DPA; the top 40 upregulated genes per compartment were classified as shared or compartment-specific. Cross-compartment ligand–receptor trafficking interactions were assessed over a curated panel of 12 LR pairs (including CCL2/CCR2, CX3CL1/CX3CR1, CSF1/CSF1R); LR scores were defined as the arithmetic mean of mean ligand expression in the sending and mean receptor expression in the receiving compartment on log-normalized data.

#### RNA velocity

RNA velocity was estimated under the dynamical model implemented in scVelo v0.3.3^30^, yielding per-cell velocity vectors, first- and second-moment matrices, and per-gene kinetic parameters (transcription, splicing, and degradation rates). Velocity streamlines were projected onto the shared UMAP embedding of integrated myeloid cells. Diffusion pseudotime (DPT) was computed using scanpy.tl.dpt with the root anchored in PBMC myeloid cells, treating blood as the origin state of a putative blood-to-lung differentiation trajectory.

#### Cell–cell communication analysis

Intercellular communication was assessed with NicheNet^31^ (v2.2.1.1) and LIANA^32^ (v0.1.9) using default prior networks and significance criteria. For NicheNet analyses, ciliated and secretory epithelial cells were defined as senders and monocyte/macrophage subpopulations as receivers; ligand activity was ranked by AUPR (corrected Pearson correlation between predicted and observed target gene expression). Differential communication networks between conditions (RDS 0–1 vs. RDS grade >=2 (2+); very preterm vs. extreme preterm) were computed as the difference in communication scores between groups.

#### Circulating epithelial cell mapping

To assign reference identities to rare epithelial cells detected in the PBMC fraction, we applied scSimilarity^33^, which maps query cells against a reference corpus of >25 million single-cell transcriptomes from the CZ CELLxGENE Discover database. For each query cell, Pearson correlation between the query and reference cell embeddings was used to identify the most similar reference cells. Cluster-level identity was assigned based on the predominant cell-type annotation of the matched reference cells, enabling annotation of circulating airway epithelial populations by their transcriptional similarity to tissue-resident counterparts spanning diverse disease states including COVID-19, IPF, COPD, and chronic rhinosinusitis.

#### Marker validation

A composite ciliated-cell module score was computed from canonical motile-cilia markers (EPCAM, FOXJ1, TPPP3, TUBB4B, DNAH5, DNAH11, RSPH1, RSPH4A, HYDIN, SPEF2, PIFO, SNTN, CDC20B, DEUP1) using scanpy.tl.score_genes. Feature plots and module scores were projected onto the integrated UMAP.

### Proteome analysis

#### Sample preparation

One µL of plasma or DPA was added to with 24 µL of lysis buffer (0.01% DDM, 10 mM Tris(2-carboxyethyl)phosphine (TCEP), 40 mM CAA in 100 mM Tris-HCl, pH 8.5) in an Eppendorf twin.tec 96-well loBind plate. Denaturation, reduction and alkylation was done by heat at 90 °C in a PCR cycler. After cooling to room temperature Trypsin and Lys-C in ratio of 1:100 to the protein amount was added, where the protein concentration for plasma was assumed at 50 mg/ml and the Protein concentration for TA was extrapolated by nanodrop measurement after denaturation. Proteins were digested at 37 °C and 1500 rpm in a thermoshaker overnight. 500 ng of the digested Peptides were desalted and loaded onto Evosep Tips following the manufacturers Protocol.

#### LCMS Data Acquisition

Peptides were separated on an 8 cm Aurora Rapid XT UHPLC column (AUR4-80150C18-XT, IonOpticks) at 50 °C using the Evosep One system^34^ with the ‘100 samples per day’ method, utilizing pre-formed gradients with a total runtime of 11.5 minutes per sample. Mass spectrometry data were acquired on a Thermo Scientific Orbitrap Astral mass spectrometer equipped with a FAIMS (high-field asymmetric waveform ion mobility spectrometry) interface operated in standard resolution mode, with a compensation voltage (CV) of −40 V and a carrier gas flow of 3.5 L/min. Full MS1 scans were acquired over a mass range of 380–980 m/z. For data-independent acquisition (DIA) MS2 scans, the precursor mass range was divided into 150 isolation windows of 4 Th utilizing window placement optimization, fragmented with an HCD collision energy of 25%, and acquired over a fragment scan range of 150–2,000 m/z.

#### MS raw data analysis and spectral search

Raw DIA files were processed with DIA-NN v2.2.0^35^ on a high-performance computing cluster. A spectral library was predicted in silico with the DIA-NN deep-learning predictor from a human UniProt Swiss-Prot isoform database (UP000005640; 42,439 entries). Trypsin/P was specified as the protease with up to one missed cleavage; the predicted library spanned peptide lengths of 7–30 residues, precursor charges of 1–4, precursor m/z of 300–1,800, and fragment m/z of 200–1,800. Carbamidomethylation of cysteine was set as a fixed modification; methionine oxidation and protein N-terminal acetylation as variable modifications (maximum one variable modification per peptide). Match-between-runs was enabled (--use-quant and --reanalyse) with peak centering, smart profiling, retention-time profiling, and relaxed protein inference. Mass accuracy was set to 10 ppm for MS1 and MS2 and the scan window to 7; cross-run normalization used the default RT-dependent strategy. Protein quantification used MS1 and MS2 without interference signal removal. The false-discovery rate was controlled at 1% at the precursor level. Quantitative matrices were used for downstream statistical analyses as described below.

#### Protein-group matrix processing

Proteome analyses were performed in R v4.5.0 and Perseus v2.1.4^36^. A fixed random seed (42) was set for all stochastic steps, and package versions were pinned with renv to ensure reproducibility. Plasma and DPA samples were processed and analysed independently throughout.

Protein-group intensities from the DIA-NN pg_matrix output were log₂-transformed; zero values were treated as missing. No additional normalization was applied, as DIA-NN MaxLFQ intensities are already cross-run normalized. Within each sample type, protein groups were retained for analysis if they were quantified (non-missing) in at least 60% of samples in at least one BPD-severity stratum (non-BPD, grade 1, or grades 2–3 combined).

Missing values were assumed to be predominantly left-censored (missing-not-at-random). They were imputed per sample from a down-shifted, width-reduced Gaussian using DEP::impute(fun = “man”)^37^ with default parameters (down-shift = 1.8 SD, width = 0.3 SD of each sample’s observed log₂ intensities). Protein groups that were entirely undetected within one BPD stratum but present in another (“on/off” proteins) were flagged, since their imputed values—and therefore any apparent between-group difference—are determined by the imputation rather than measurement; these were interpreted as qualitative detections rather than graded changes.

#### Correlation with gestational age

Within each sample type (plasma and DPA), filtered and imputed log2 protein intensities were correlated with gestational age (weeks at birth) using Spearman rank correlation per protein. Proteins reported as ‘GA-associated’ (Fig. 5B) had |Spearman ρ| > 0.5 with respect to gestational age. P-values were BH-adjusted within each sample type.

#### Functional enrichment analysis

Protein enrichment analysis was assessed with clusterProfiler(4.16.0)^38^ using over-representation analysis (ORA). Gene sets comprised Gene Ontology Biological Process, Cellular Component and Molecular Function (org.Hs.eg.db 3.21.0), Reactome (msigdbr v25.1.1), and UniProt Keywords (retrieved UniProt API (release 2026_02)). Gene symbols were taken from the leading protein of each group. The background universe for each test was the set of proteins quantified in that tissue. p-values were BH-adjusted within each collection.

#### Organ-to-System Score

For each donor, the Organ-to-System Score was computed as the weighted sum of log₂-transformed plasma intensities of lung-restricted marker proteins detected in blood. Proteins were assigned weights according to tissue specificity: ciliated airway epithelial cell markers received weight 3×, reflecting their near-exclusive expression in the airway epithelium under physiological conditions; alveolar type II (AT2) cell markers received weight 2×; remaining lung-associated markers received weight 1×. This weighting scheme ensures that the score preferentially reflects the presence of the most lung-restricted proteins in the systemic circulation, providing a sensitive readout of lung epithelial barrier disruption.

#### Disease trajectory analysis

Preterm Donors (<32 weeks) were classified into three disease trajectory groups based on combined RDS and BPD severity scores: Healthy (RDS 0–1, BPD 0–1; n = 5), Acute-only (RDS grade >=2 (2+), BPD degree 0–1; n = 7), and Acute-to-chronic (RDS 2+, BPD degree moderate (2) and severe (3) (2+); n = 5). For each plasma protein passing detection and variance filters, Spearman ρ between protein abundance and BPD severity and between protein abundance and RDS severity was computed across all plasma samples. Trajectory-specific proteins were identified by one-versus-rest Spearman correlation (ρ ≥ 0.5) between protein abundance and group membership. Module scores from trajectory-specific protein signatures were projected onto DPA and PBMC scRNA-seq datasets using scanpy.tl.score_genes. Pathway enrichment of trajectory-specific signatures was performed as described above (ORA, clusterProfiler).

### Isolation of mononuclear cells and flow cytometry

Mononuclear cells were isolated from EDTA-supplemented umbilical cord blood samples using density-gradient centrifugation using Ficoll Paque Premium (Thermo Fisher Scientific) according to standard methods. Following centrifugation, the mononuclear cell fraction was collected and washed with PBS to remove residual plasma, platelets, and density-gradient medium. Subsequently, cells were resuspended in an appropriate buffer and cell number (CASY cell counting, Omni Life Sciences) and viability were determined using the Pacific Blue™ Annexin V Apoptosis Detection Kit with PI (Biolegend, San Diego, CA) and CellEvent™ Caspase-3/7 Detection Reagent in combination with the SYTOX Advanced Dead Cell Staining Kit Thermo Fisher Scientific) according to the manufacturer’s instructions. Isolated mononuclear cells were either processed immediately for downstream multicolor flow cytometry, or cryopreserved. All samples were handled in accordance with institutional ethical approvals and biosafety regulations for human biological material and experiments were carried out according to the Declaration of Helsinki.

The expression of cell surface and intracellular markers was analyzed by multicolor flow cytometry on a CytoFLEX LX machine (Beckman Coulter) using the CytExpert (acquisition) and Kaluza software (analysis). For flow cytometry, cells were stained in 50 µl of PBS using appropriate dilutions of antibodies against BV650-coupled anti-human CD14 (clone M5E2), PE-coupled anti-human CD16 (clone 3G8), BV650-coupled anti-human CD45 (clone HI30), PE-Dazzle594-coupled anti-human CD207 (clone 4C7), BV650-coupled anti-human CD326 (clone 9C4), PerCP-Cy5.5-coupled anti-human CD68 (clone Y1/82A), AF700-coupled anti-human HLA-DR (clone L243), BV785-coupled anti-human CD11b (clone M1/70), BV421-coupled anti-human CD31 (clone WM59), PerCP-Cy5.5-coupled anti-human CD195 (clone J418F1), APC-coupled anti-human CD186 (clone KO41E5), FITC-coupled anti-human CD197 (clone GO43H7), APC-Cy7-coupled anti-human CD199 (clone LO53E8), BV510-coupled anti-human CD183 (clone GO25H7), AF700-coupled anti-human CD1a (clone HI149), PE-coupled anti-human CD184 (clone QA18A64), PE-Cy7-coupled anti-human CD185 (clone J252D4), BV421-coupled anti-human IL-6 (clone MQ2-13A5; all purchased from BioLegend). Intracellular staining was performed after cell permeabilization using the Fix/Perm Buffer Set (BioLegend) according to the manufacturer’s instructions. Isotype-matched controls were included in each staining and statistical evaluation to compare data from pre-term versus full-term infants was calculated using the Student’s t-test and and a *p*-value < 0.05 was considered as statistically significantly different. Flow cytometry was performed using 2 different panels with panel A containing antibodies to CD68, CD16, HAL-DR, IL-6, CD45, CD14, CD11b and panel B containing antibodies to CD197, CD195, CD184, CD207, CD185, CD186, CD1a, CD199, CD31, CD183, CD45, CD326. Prior to flow cytometric acquisition, spectral overlap was corrected by generating a compensation matrix using UltraComp eBeads (Thermo Fisher Scientific) according to the manufacturer’s protocol. In addition, unstained cells were included during compensation setup to account for cell-intrinsic autofluorescence and to reduce false-positive signals within the analyzed cell populations.

#### Flow cytometric gating strategy for macrophages, monocytes, and epithelial cells

For the analysis of macrophages and monocytes, dead cells were excluded prior to gating. Subsequently, leukocytes were identified by gating on CD45⁺ cells, followed by singlet discrimination using SSC-H versus SSC-A. For macrophage analysis, the relative frequency of CD68⁺ cells was determined, with CD68 serving as a lineage marker for predominantly pro-inflammatory macrophages. CD11b was intentionally not included in the gating strategy, as its expression can be downregulated in specific macrophage subsets, such as alveolar macrophages, and may further vary depending on the activation state. To assess macrophage activation, the proportion of IL-6⁺CD68⁺ cells was quantified as a marker of activated pro-inflammatory macrophages. Monocyte analysis was performed using a comparable gating strategy. Following gating on viable CD45⁺ singlet cells, the relative frequencies of CD14⁺ and CD16⁺ populations were determined. Monocyte subsets were defined as CD14⁺CD16⁻ classical monocytes and CD14dimCD16⁺ non-classical monocytes. For epithelial cell analysis, viable CD45⁻ singlet cells were selected first. Within this population, epithelial cells were defined as CD1a⁻CD207⁻CD326⁺ cells and used for downstream analyses. Exclusion of CD1a⁺CD207⁺ cells was necessary because certain dendritic cell subsets, including epidermal Langerhans cells, are known to express CD326 and could otherwise confound epithelial cell identification. Within the CD45⁻CD1a⁻CD207⁻CD326⁺ population, the relative frequencies of cells expressing specific chemokine receptors associated with migratory behavior or tissue tropism were quantified. These included CD183 and CD186 (lung-associated homing), CD197 (lymph node homing), CD199 (gut-associated homing), as well as CD184 and CD185, which have been associated with trafficking to the central nervous system.

### Statistical analysis

Statistical analyses of flow cytometry data were performed using GraphPad Prism (version 9). Relative frequencies of CD68⁺ macrophages in preterm and full-term infants (Figure 2A) were compared using an unpaired two-tailed Student’s *t*-test, as two independent experimental groups were analyzed. The same statistical approach was applied to the analysis of CD68⁺IL-6⁺ macrophages and monocyte subsets (Figure 2A). Sample sizes were *n* = 31 for CD68⁺ cells and monocytes in preterm infants and *n* = 28 for CD68⁺ cells and monocytes in full-term infants. For CD68⁺IL-6⁺ cells, sample sizes were *n* = 17 in preterm infants and *n* = 16 in full-term infants. Statistical analysis of CD326⁺ epithelial cells in cord blood (Figure 3A) was likewise performed using an unpaired two-tailed Student’s *t*-test (*n* =31 preterm infants; *n* = 28 full-term infants). Differences in the relative frequencies of epithelial cells expressing distinct chemokine receptors were analyzed using two-way analysis of variance (two-way ANOVA), with gestational age (preterm vs. full-term) and chemokine receptor expression as independent variables in *n* = 20 preterm infants. Where appropriate, multiple comparisons were corrected using a Sidak’s test. All flow cytometric data are presented as mean ± standard deviation (SD). A *p* value < 0.05 was considered statistically significant and is indicated by an asterisk (*).

All other statistical analyses were performed in R v4.4 or Python 3. Group comparisons for continuous variables were performed using the Wilcoxon rank-sum test. Correlation analyses used Spearman’s rank correlation. For analyses involving repeated measures from the same donor, linear mixed models (LMM) were fitted using the lme4 package in R with donor identity as a random intercept. Multiple testing correction was applied using the Benjamini–Hochberg (BH) procedure throughout; adjusted p-values < 0.05 were considered statistically significant unless stated otherwise.

Portions of the analysis code (R and Python scripts) were drafted with the assistance of Claude (Opus 5, Anthropic). All AI-generated code was reviewed, executed and validated by the authors.

## Results

### Gestational immaturity reshapes airway and blood immune landscapes

To define coordinated immune adaptation across the airway–blood axis, we profiled paired airway (DPA) and blood (PBMC) samples from neonates spanning extreme prematurity to term gestation at two early postnatal timepoints (timepoint 1 (day of life 1-3) and timepoint 2 (day of life 4-10) (**Fig. 1A**; **Table 1; Supplementary Table 1**). The cohort spanned a broad spectrum of gestational maturity and corresponding respiratory morbidity, with increasing prematurity associated with a greater need for respiratory support and acute respiratory disease (RDS), development of prematurity-associated chronic lung disease (BPD, bronchopulmonary dysplasia), and prolonged hospitalization (**Table 1**).

**Figure 1.**
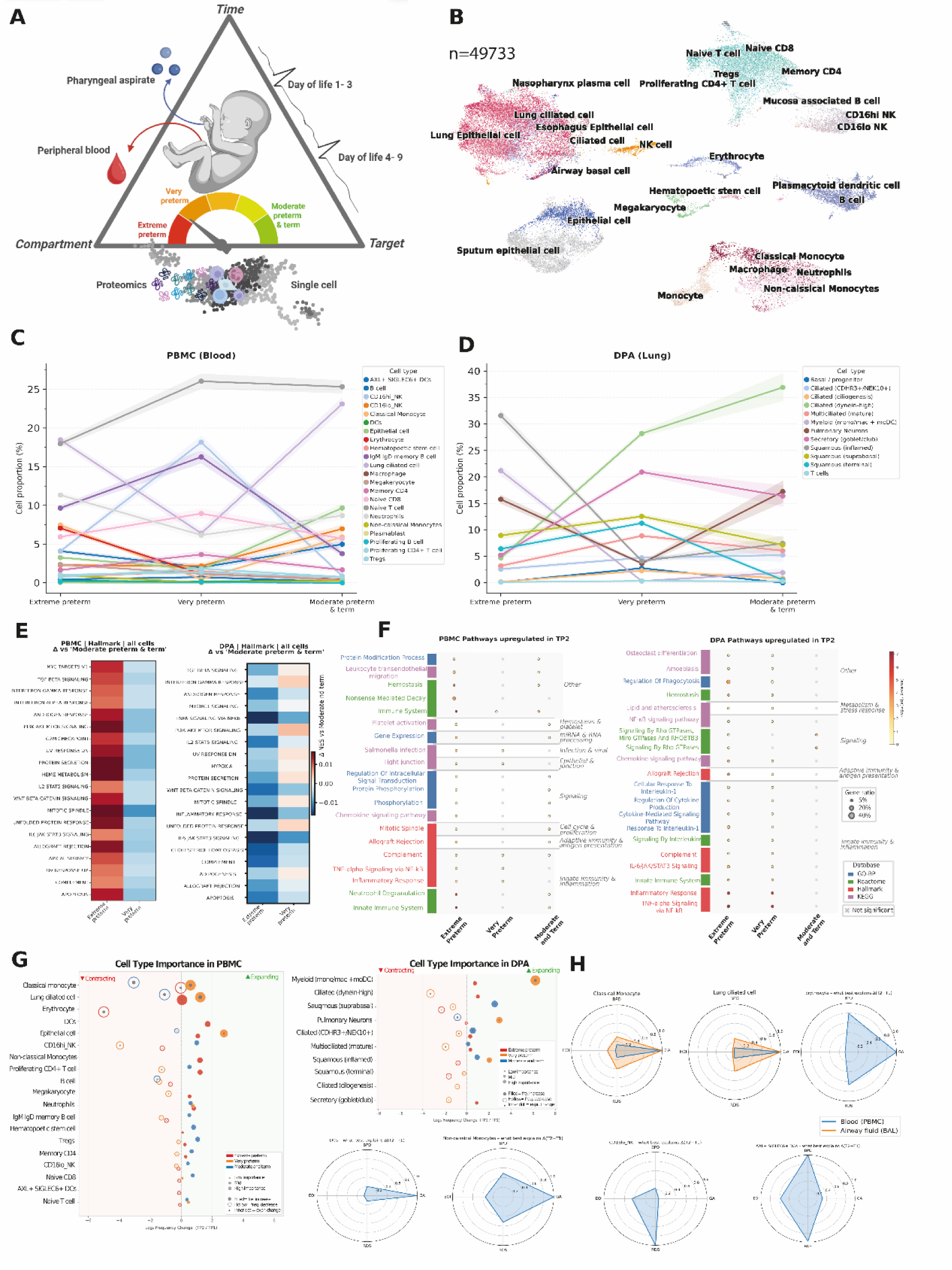
Gestational immaturity drives coordinated yet compartment-specific immune remodeling across the airway–blood axis during early postnatal life. **(A)** Study design. Paired DPA and blood samples were collected from neonates across three gestational age groups (extreme preterm <28 weeks; very preterm 28–32 weeks; moderate preterm/term ≥32 weeks) at two postnatal timepoints (T1: day of life 1–3; T2: day of life 4–10) for single-cell RNA sequencing (cell pellet) and proteomics (supernatant). Created with biorender.com. **(B)** Integrated UMAP of single-cell transcriptomes from merged DPA and PBMC samples (N=49733) across both timepoints, colored by annotated cell type (monocytes, macrophages, DC, NK, T, B, erythroid/HSC, megakaryocytes, ciliated, secretory, basal, squamous epithelial). **(C, D)** Relative cell-type proportions across gestational age groups in PBMC **(C)** and DPA **(D)** at the early postnatal timepoint, showing compartment-specific shifts in immune and epithelial composition. **(E)** Hallmark pathway enrichment heatmaps showing pathways differentially regulated in extreme preterm and very preterm neonates relative to moderate preterm/term neonates in PBMC and DPA at the early postnatal timepoint. Red indicates higher enrichment in the preterm group; blue indicates lower. **(F)** Dot plots of pathways upregulated at the late postnatal timepoint in PBMC and DPA, stratified by gestational age group and grouped by biological process and database source. **(G)** Cell-type importance analysis showing expanding and contracting populations across postnatal time in PBMC and DPA. **(H)** Radar plots of compartment- and cell-type-specific functional signatures associated with gestational age, lung morbidity (RDS, BPD), and early-onset infection. *Abbreviations:* PBMC, peripheral blood mononuclear cell; DPA, deep pharyngeal aspirate; NK, natural killer cell; DC(s), dendritic cell(s); moDC, monocyte-derived dendritic cell; Tregs, regulatory T cells; CD4/CD8, cluster of differentiation 4/8; CD16hi/lo, CD16-high/low; GA, gestational age; TP1/TP2, timepoint 1/timepoint 2 (day of life 1–3 / day of life 4–10); NES, normalized enrichment score; ssGSEA, single-sample gene set enrichment analysis; GO-BP, Gene Ontology Biological Process; KEGG, Kyoto Encyclopedia of Genes and Genomes; UMAP, Uniform Manifold Approximation and Projection; EOI, early-onset infection; RDS, respiratory distress syndrome; BPD, bronchopulmonary dysplasia.

After quality control, 49,733 high-quality cells from paired airway (15514) and blood (34219) samples were retained for downstream analyses (**Supplementary Fig. S1**). Integrated single-cell profiling established a cellular atlas of the neonatal airway–blood axis comprising immune, hematopoietic, and epithelial populations across paired airway and blood compartments **(Fig. 1B)**: classical and non-classical monocytes, macrophages, dendritic cells, NK cells, T and B cells, erythroid and hematopoietic progenitors, and ciliated, secretory, basal, and squamous epithelial cells. The presence of both immune and epithelial populations established a framework to define coordinated and compartment-specific immune adaptation across the airway–blood axis during the first week of life. The data are available for interactive exploration at CELLxGENE (<u>link</u>).

Across maturity groups, PBMCs showed gestational age-associated shifts in immune composition during the first 72 hours of life **(Fig. 1C)**. Variations in naive T cells, NK cell subsets, monocytes, and erythroid/hematopoietic populations, consistent with systemic immune composition, are already shaped by gestational maturity at birth. Notably, epithelial-associated cells were also detected in the PBMC compartment, an observation we revisit in detail below. In airway samples, epithelial composition varied more prominently across maturity groups, with increasing representation of ciliated and secretory epithelial cells in more mature infants, alongside shifts in myeloid and squamous epithelial populations **(Fig. 1D)**. These findings indicate that developmental maturity drives coordinated remodeling across the airway–blood axis while preserving compartment-specific cellular programs.

Pathway analysis revealed that these compositional differences were accompanied by compartment-specific patterns of immune activation. In PBMCs, extreme preterm infants showed higher enrichment of inflammatory, interferon, STAT-associated, and stress-response pathways compared with moderate preterm/term infants **(Fig. 1E)**. In DPA, the pattern was distinct: extreme preterm infants showed reduced activation of several pathways and a relative shift of interferon-associated activity toward very preterm infants. This divergence indicates that gestational maturity does not impose a uniform immune state across compartments, but differentially affects systemic and airway-localized responses during early postnatal life. These findings demonstrate that developmental maturity coordinately remodels the airway–blood axis while differentially shaping immune activation within each compartment.

At the later postnatal timepoint, pathways upregulated in PBMC and DPA indicated persistent innate immune and inflammatory activity, particularly in infants with significant immaturity **(Fig. 1F)**. These data are consistent with immune activation during the first week of life evolving across both systemic and airway compartments rather than being confined to the immediate postnatal transition.

Cell-type importance analysis identified monocytes and myeloid populations as major contributors to postnatal remodeling in blood and DPA, respectively **(Fig. 1G)**. Epithelial populations also emerged as important maturity- and time-associated features, supporting a model in which epithelial maturation and immune adaptation are linked during early life rather than reflecting separable processes. Radar plots highlighted compartment- and cell-type-specific functional signatures associated with gestational age, lung morbidity (RDS and BPD), and early-onset infection (EOI) (**Fig. 1H**). Classical and non-classical monocytes, together with epithelial populations, contributed prominently to maturity- and lung disease-associated variation, whereas NK and dendritic cell signatures were more strongly associated with infection-related variation. Rather than a uniform program of immune immaturity, these findings reveal coordinated yet compartment-specific remodeling across the airway–blood axis, with epithelial and myeloid programs emerging as the principal drivers of early postnatal immune adaptation and its disruption in extreme prematurity.

#### Myeloid remodeling links prematurity to neonatal lung disease

Since we identified monocyte and myeloid populations as major contributors to maturity- and disease-associated variation, we next focused on myeloid remodeling across blood and airway compartments. Flow cytometry of peripheral blood *CD45*+ cells confirmed altered myeloid composition in preterm infants (n=31, GA 25-35 weeks) compared with full-term neonates (n=28, GA 38-40 weeks), including reduced *CD14*+*CD16*-classical monocytes, increased *CD14*dim*CD16*+ non-classical monocytes, and reduced frequencies of CD68+ macrophages and *CD68*+*IL-6*+ pro-inflammatory macrophages (**Fig. 2A**). These flow cytometry findings indicate that gestational immaturity is associated with measurable shifts in circulating myeloid composition and activation state.

**Figure 2.**
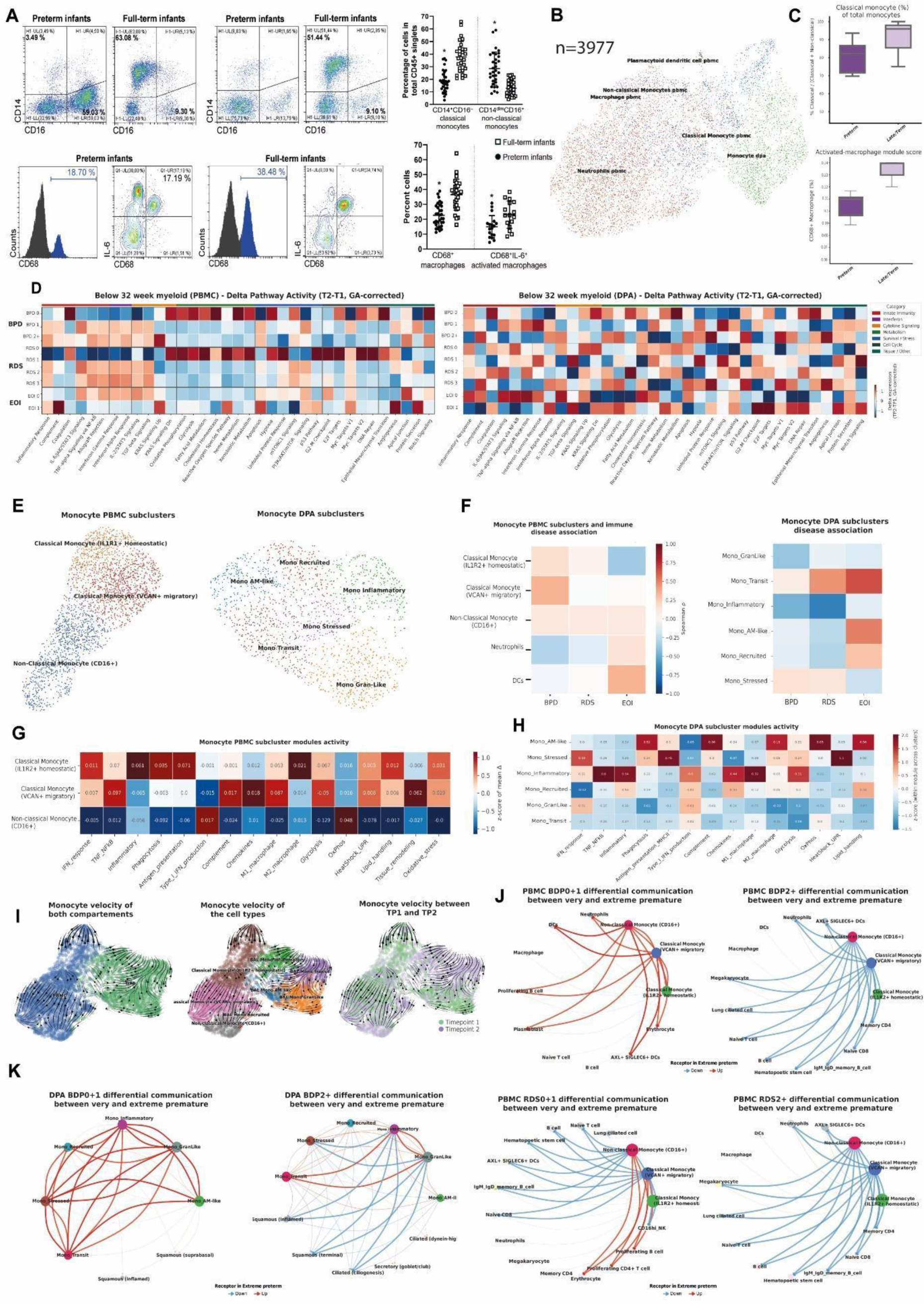
Myeloid remodeling coordinates immune adaptation across the airway–blood axis and associates with neonatal lung disease. **(A)** Flow cytometry of CD45+ singlets from cord blood mononuclear cells. Pseudocolor plots show CD14 vs CD16 staining to distinguish classical (CD14+CD16-) from non-classical (CD14dimCD16++) monocytes and to assess the relative numbers of total CD68+ macrophages as well as CD68+IL-6+ activated, pro-inflammatory macrophages. Scatter plots show quantification across preterm and term groups. **(B)** UMAP of n = 3,977 myeloid cells from integrated PBMC and DPA, annotated by cell type (classical monocytes, non-classical monocytes, macrophages, pDCs, neutrophils, DPA monocyte/macrophage states). **(C)** Box plots of classical monocyte fraction (top) and activated macrophage module score in CD68+ macrophages (bottom), comparing preterm and late preterm/term infants. P-values from two-sided Wilcoxon rank-sum tests; BH-adjusted within the comparison family. **(D)** Heatmaps of gestational age-corrected ΔT2–T1 pathway activity in PBMC and DPA myeloid cells, stratified by BPD, RDS, and EOI severity. Color encodes the LMM β coefficient; asterisks mark BH-adjusted q < 0.05. **(E)** UMAP projections of monocyte subclusters in PBMC (left) and DPA (right). PBMC: IL1R2+ homeostatic classical, VCAN+ migratory classical, CD16+ non-classical. DPA: Mono_Recruited, Mono_Inflammatory, Mono_AM-like, Mono_Stressed, Mono_Transit, Mono_GranLike. **(F)** Heatmap of functional module scores across PBMC monocyte subclusters (IFN response, TNF–NF-κB, phagocytosis, antigen presentation, complement, chemokines, M1/M2 polarization, glycolysis, oxidative phosphorylation, lipid handling, tissue remodeling, oxidative stress). Z-scored mean module activity per subcluster. **(G)** Equivalent module-score heatmap across DPA monocyte/macrophage subclusters. **(H)** Disease-association heatmap (Spearman ρ between subcluster abundance and BPD, RDS, EOI severity) for PBMC (left) and DPA (right) myeloid subclusters; asterisks denote nominal p < 0.05. **(I)** RNA velocity streamline plots of monocytes from both compartments combined, annotated by cell type, and colored by timepoint (TP1, green; TP2, blue). Arrows indicate predicted differentiation trajectories. **(J)** Communication plots of IL1R2_homeostatic, VCAN_migratory, and CD16_NCM module scores across PBMC monocyte subclusters and DPA monocyte/macrophage populations. **(K)** Differential intercellular communication networks comparing very preterm and extreme preterm infants across disease strata. Networks show differential ligand–receptor signaling involving myeloid cells; red edges = increased signaling in extreme preterm, blue = decreased. Edge thickness reflects communication strength. *Abbreviations:* PBMC, peripheral blood mononuclear cell; DPA, distal pharyngeal aspirate; AM, alveolar macrophage; DC(s), dendritic cell(s); CD4/CD8/CD14/CD16/CD68, cluster of differentiation 4/8/14/16/68; IL-6/IL2R1/IL2R2, interleukin-6/interleukin-2 receptor 1/2; VCAN, versican; TNF, tumor necrosis factor; NF-κB, nuclear factor kappa-B; IFN, interferon; OxPhos, oxidative phosphorylation; UPR, unfolded protein response; GA, gestational age; TP1/TP2, timepoint 1/timepoint 2 (day of life 1–3 / day of life 4–10); BPD, bronchopulmonary dysplasia; RDS, respiratory distress syndrome; EOI, early-onset infection; UMAP, Uniform Manifold Approximation and Projection.

Single-cell analysis of integrated PBMC- and deep pharyngeal aspirate-derived myeloid cells identified 3,977 myeloid cells across compartments, including classical monocytes, non-classical monocytes, macrophages, plasmacytoid dendritic cells, neutrophils, and airway-enriched monocyte/macrophage populations (**Fig. 2B; Fig. S2A–C)**. Quantification of myeloid features confirmed differences in classical monocyte representation and activated-macrophage module activity between preterm and more mature infants (**Fig. 2C**), consistent with the flow cytometry results and extending these observations at single-cell resolution.

To determine whether myeloid remodeling was linked to neonatal respiratory disease, we next examined pathway activity across clinical phenotypes. PBMC monocytes from preterm infants carried gestational age- and disease-associated changes in interferon, *TNF–NF-κB*, and chemokine interaction signatures (**Fig. 2D**). In airway-derived myeloid cells, pathway changes were more heterogeneous across lung (RDS, BPD), and systemic infection (EOI) strata, consistent with stronger compartment-specific priming or local adaptation within the airway environment (**Fig. 2D**).

Subclustering revealed discrete circulating and airway myeloid states that capture progressive differentiation and tissue adaptation across the airway–blood axis **(Fig. 2E)**. PBMC monocytes included *IL1R2*+ homeostatic classical monocytes, VCAN+ migratory classical monocytes, and *CD16*+ non-classical monocytes. Airway-derived monocyte/macrophage populations included recruited, inflammatory, alveolar macrophage-like, stressed, transitional, and granulocyte-like states. Disease-association analysis linked distinct myeloid states to neonatal disease features. In PBMCs, *VCAN*+ migratory classical monocytes showed association with bronchopulmonary dysplasia, while dendritic cell-associated signatures were more strongly linked to early-onset infection (**Fig. 2F**). In airway-derived myeloid cells, disease associations were distributed across inflammatory, alveolar macrophage-like, recruited, stressed, transitional, and granulocyte-like states, consistent with a more heterogeneous local myeloid response to pulmonary injury and infection **(Fig. 2F)**. Thus, neonatal respiratory disease reflects compartment-specific remodeling of multiple myeloid states rather than expansion of a single inflammatory population.

Functional module scoring showed that these subclusters differed in interferon response, TNF–NF-κB signaling, phagocytosis, antigen presentation, complement activation, chemokine signaling, macrophage polarization, metabolism, tissue remodeling, and oxidative stress programs (**Fig. 2G,H; Fig. S2D**). These programs reveal functional specialization rather than a single activated myeloid state.

To investigate the directionality of myeloid state transitions, we reconstructed transcriptional trajectories using RNA velocity across both compartments (**Fig. 2I**). In PBMCs, velocity fields supported a directional continuum from IL1R2⁺ homeostatic classical monocytes through VCAN⁺ migratory monocytes toward CD16⁺ non-classical monocytes. Within the DPA compartment, velocity vectors indicated directed transitions from recruited monocyte states toward AM-like and stressed monocyte programs, consistent with in situ maturation rather than independent differentiation. Temporal comparison between timepoints 1 (day of life 1-3) and 2 (day of life 4-10) revealed a shift in velocity field orientation, suggesting that myeloid trajectory dynamics evolve over the early postnatal period. To characterize how disease severity reshapes intercellular signaling within these myeloid networks, we performed differential cell-cell communication analysis comparing very preterm and extreme preterm infants across disease conditions (**Fig. 2J–K**). In the PBMC compartment, CD16⁺ non-classical monocytes emerged as a central communication hub, with BPD-associated upregulation of outgoing signals in extreme preterm infants concentrated in BPD grade moderate and severe (2+) (**Fig. 2J**). In the DPA compartment, Mono_Recruited and Mono_GranLike subtypes showed the most pronounced rewiring of communication networks with increasing disease severity, with BPD grade moderate and severe (2+) associated with a shift from Mono_Inflammatory-centered toward Mono_Recruited-centered outgoing signaling (**Fig. 2K**, left panels). RDS severity was associated with analogous rewiring in the PBMC compartment, with RDS grade >=2 (2+) driving broader engagement of classical monocyte subtypes as communication senders (**Fig. 2K**, right panels). These analyses reveal that disease severity does not uniformly amplify myeloid signaling but selectively restructures intercellular communication topology in a compartment- and condition-specific manner.

### Circulating respiratory epithelial-like cells are enriched in preterm blood and associate with neonatal lung disease

Our integrated paired airway–blood single-cell atlas (**Fig. 1B,C**) unexpectedly identified epithelial-like cells within the PBMC compartment. This observation prompted us to determine whether these epithelial-like cells represented a biologically meaningful population associated with developmental immaturity and neonatal lung disease rather than technical artifact. We first asked whether CD326/EPCAM+ cells could be detected in neonatal blood by flow cytometry. Gating on CD45-events and excluding rare APC subsets known to co-express CD326 (CD45-CD207-CD1a-CD326+) identified EPCAM+ cells in peripheral blood from both term and preterm infants, with higher abundance in preterm infants (**Fig. 3A**). Chemokine receptor profiling within the CD326+ gate further suggested tissue-origin heterogeneity, including marker combinations consistent with lung-associated (CD183+CD186+), lymph node-associated (CD197+), brain-associated (CD184+CD185+), and gut-associated (CD199+) epithelial-like populations (**Fig. 3B**). Cells with a lung-associated chemokine-receptor profile accounted for a substantial fraction, supporting the hypothesis that respiratory epithelial-like cells contribute to the circulating EPCAM+ signal in preterm infants.

**Figure 3.**
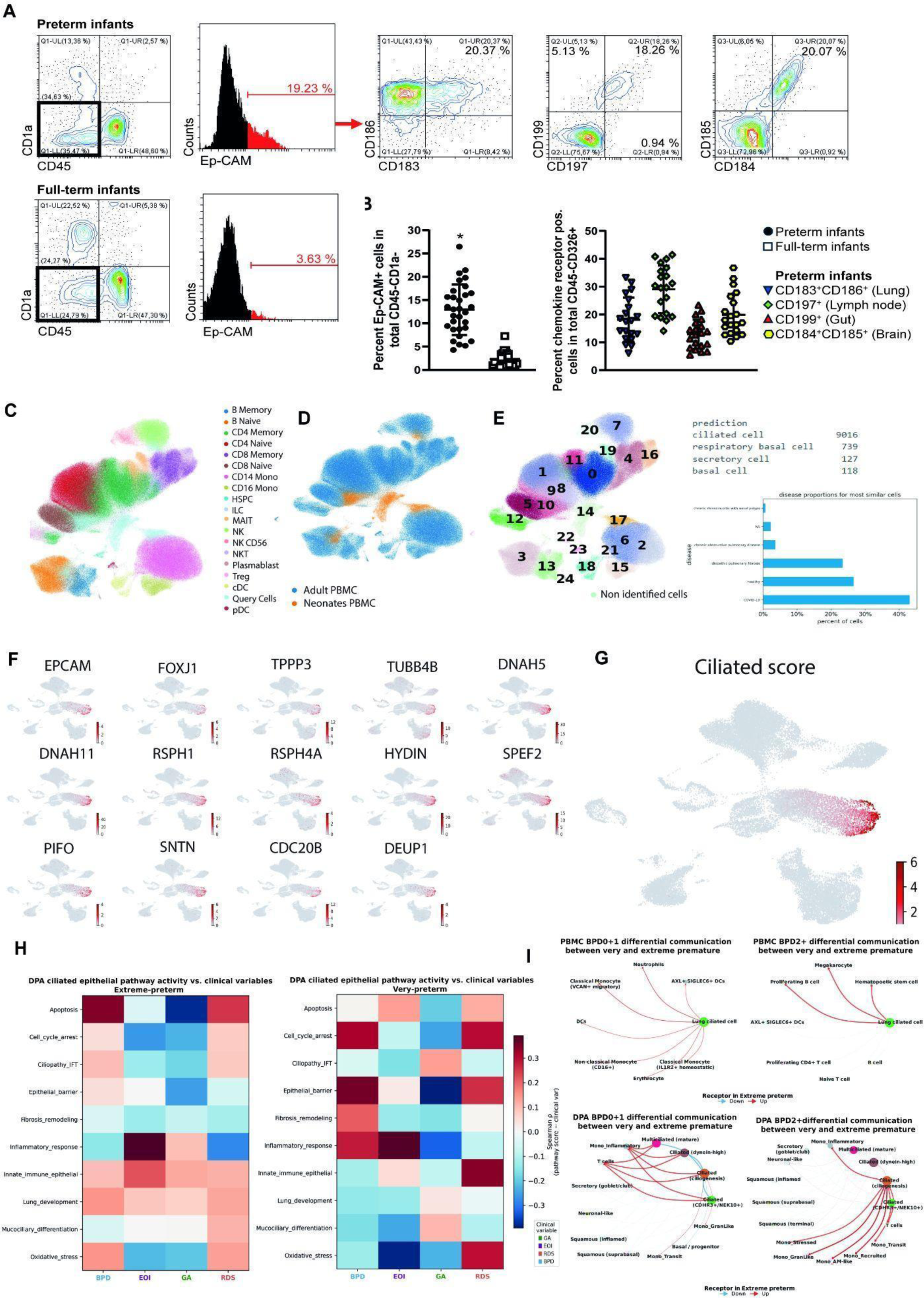
Lung epithelial cells circulate in the peripheral blood of preterm neonates and engage in disease-associated cellular crosstalk. **(A,B)** Representative flow cytometry gating strategy identifying circulating EpCAM⁺ cells within the CD45⁻CD1a⁻CD207⁻ population in term and preterm peripheral blood. The precise gating strategy is described in the material and methods section. The EpCAM⁺ population was further profiled for chemokine receptor expression (CD183/CD186, CD197, CD199, CD184/CD185 as indicated in the figure) to define tissue-homing subsets. Representative contour or histogram plots **(A)** and the statistical evaluation **(B)** are shown and each point represents one donor. Data are presented as mean±SD; *p < 0.05 and detailed information concerning sample size as well as statistical methods can be found in the material and methods section. **(C)** UMAP embedding of PBMC scRNA-seq reference data colored by canonical immune cell-type annotation (18 populations, including the projected query cells). **(D)** Same UMAP embedding as (B), colored by dataset of origin — adult PBMC reference (blue) versus neonatal PBMC query cells (orange) — showing the region of transcriptional space occupied by the neonatal circulating epithelial-like population. **(E)** *Left*, unsupervised clustering (clusters 0–24) of the query cell population, with unidentified cells indicated separately. *Right*, predicted cell identity of the dominant cluster (top: ciliated cell, n = 9,016; respiratory basal cell, n = 739; secretory cell, n = 127; basal cell, n = 118) and the disease-state composition of the most transcriptionally similar reference cells (percent of cells matching COVID-19, healthy, idiopathic pulmonary fibrosis, chronic obstructive pulmonary disease, and chronic rhinosinusitis-with-nasal-polyps reference atlases). **(F)** Feature plots of canonical ciliated-cell markers (*EPCAM, FOXJ1, TPPP3, TUBB4B, DNAH5, DNAH11, RSPH1, RSPH4A, HYDIN, SPEF2, PIFO, SNTN, CDC20B, DEUP1*) projected onto the UMAP embedding, confirming ciliogenesis/motile-cilia identity of the dominant query cluster. **(G)** Composite ciliated-cell module score projected onto the UMAP embedding, highlighting the cluster of circulating cells with a transcriptional signature consistent with airway ciliated epithelium. **(H)** Heatmaps of Spearman correlation (ρ) between deep pharyngeal aspirate (DPA) ciliated-epithelial pathway activity scores (rows; ten curated gene-set modules spanning apoptosis, cell-cycle arrest, ciliopathy/intraflagellar transport, epithelial barrier, fibrosis/remodeling, inflammatory response, innate immune function, lung development, mucociliary differentiation, and oxidative stress) and clinical variables (columns: BPD, EOI, GA, RDS), shown separately for extreme-preterm (left) and very-preterm (right) donor subgroups. Color scale indicates Spearman ρ; clinical variables are color-coded per the legend. **(I)** Cell– cell communication network plots showing differential ligand–receptor signaling between very-preterm and extreme-preterm donors, stratified by BPD severity (BPD0+1 vs. BPD2+) and compartment (PBMC, top; DPA, bottom). Nodes represent cell types (lung ciliated cells as the central hub in PBMC; ciliated, secretory, basal, and monocyte subsets in DPA); edge color indicates the direction of receptor expression change in extreme-preterm relative to very-preterm donors (red = up, blue = down). *Abbreviations:* PBMC, peripheral blood mononuclear cell; DPA, distal pulmonary aspirate; (CD326 = EpCAM); EpCAM, epithelial cell adhesion molecule; CCR5/CCR6/CCR7/CCR9, C-C chemokine receptor type 5/6/7/9; CXCR3, C-X-C chemokine receptor type 3; HSPC, hematopoietic stem and progenitor cell; ILC, innate lymphoid cell; MAIT, mucosal-associated invariant T cell; NK, natural killer cell; NKT, natural killer T cell; Treg, regulatory T cell; cDC/pDC, conventional/plasmacytoid dendritic cell; Mono, monocyte; IFT, intraflagellar transport; GA, gestational age; BPD, bronchopulmonary dysplasia; RDS, respiratory distress syndrome; EOI, early-onset infection; UMAP, Uniform Manifold Approximation and Projection.

To further define the identity of these cells, we integrated neonatal PBMCs with an adult PBMC reference atlas, revealing a discrete neonatal-enriched cluster falling outside the adult immune space (**Fig. 3C,D**), consistent with a neonatal-associated cell state rather than a common circulating immune population. Reference-based cell-type prediction classified this population with highest confidence as ciliated respiratory epithelial cells, with smaller contributions from predicted respiratory basal and secretory identities (**Fig. 3E**). The most similar reference cells in disease-association space were drawn from idiopathic pulmonary fibrosis and COVID-19 datasets, supporting a respiratory epithelial identity distinct from canonical circulating immune populations.

To validate the epithelial and respiratory identity of this cluster, we examined canonical epithelial and ciliated-cell markers across the integrated UMAP. The epithelial marker EPCAM and the ciliated/respiratory markers *FOXJ1, TPPP3, TUBB4B, DNAH5, DNAH11, RSPH1, RSPH4A, HYDIN, SPEF2, PIFO, SNTN, CDC20B,* and *DEUP1* were enriched within the neonatal epithelial-like cluster (**Fig. 3F**). A composite ciliated-cell score localized to the same cluster (**Fig. 3G**), consistent with a coordinated ciliated respiratory epithelial transcriptional program. Given the unexpected nature of this finding, we systematically evaluated alternative technical explanations, including (i) : (i) cluster doublets between blood cells and co-processed DPA epithelial cells, (ii) ambient-RNA contamination from co-loaded libraries, and (iii) carry-over from buffy-coat or pellet handling. Standard doublet detection and ambient-RNA correction were applied (scDblFinder for doublets, SoupX for ambient RNA) and the cluster retained its coordinated EPCAM and ciliated-marker signature after correction. PBMC and DPA samples were processed and indexed independently to limit cross-compartment contamination. These observations argue against the alternative explanations and support a biologically structured population, while leaving the question of tissue origin and route of entry into the circulation.

To further strengthen the finding, re-analysis in a published PBMC single-cell RNA dataset^11^ revealed a discrete population of circulating cells expressing a canonical lung ciliated epithelial signature that was not reported in the original study. Unsupervised clustering and transcriptomic annotation identified 61 cells (0.27% of total PBMCs) characterized by high co-expression of FOXJ1, DNAH5, PIFO, TPPP3, RSPH1, RSPH4A, SPEF2, SNTN and CDC20B, markers collectively diagnostic of motile ciliated airway epithelium (**Supplementary Fig. 4**). These cells formed a transcriptionally coherent and reproducible cluster clearly distinct from all haematopoietic lineages in the dataset, suggesting they represent genuine circulating epithelial cells rather than ambient RNA contamination or multiplets. Their presence in the peripheral blood fraction raises the possibility that epithelial shedding or trafficking occurs in this context, a phenomenon that went unrecognized in the original analysis. Importantly, the independent identification of this circulating ciliated cell population in an entirely separate cohort and dataset directly corroborates our own findings, reinforcing the robustness and biological relevance of this observation across datasets.

To validate the transcriptional identity of the circulating lung ciliated cells identified in the PBMC dataset, we performed reference-based integration of these cells onto a published neonatal lung single-cell RNA-sequencing atlas^39^ (GSE275938; 67,419 cells). Using a scArches query-mapping approach, we first trained an scVI model on the neonatal lung dataset from Shiraz et al. as the reference, then projected the PBMC-derived lung ciliated cells as a query into the pre-established latent space (**Supplementary Fig. 5A–B**). In the resulting joint UMAP embedding, the PBMC ciliated cells co-localized almost exclusively with a discrete cluster of the atlas that was annotated as ciliated epithelium (Cluster 12; **Supplementary Fig. 5C**). K-nearest neighbour (KNN) analysis confirmed this spatial proximity: 58.5% of the KNN contacts of PBMC ciliated cells fell within the ciliated cluster, representing a 20.8-fold enrichment over the expected frequency based on cluster size (**Supplementary Fig. 5D, H**). The mean query ciliated neighbour score was 0.0353 for the ciliated cluster, more than 13-fold higher than the next-ranked cluster (Supplementary Figure 5G). No other major lung cell type (endothelial, fibroblast, AT2, T/NK) reached a fold-enrichment above 1.8× (Supplementary Figure 5H). Consistent with this proximity, ciliated marker genes, including FOXJ1, DNAH5, TPPP3, PIFO, SNTN, CDC20B, RSPH1, RSPH4A, SPEF2, and EPCAM, were expressed at high levels and in a high fraction of cells exclusively within the atlas ciliated cluster, with negligible expression in all other clusters examined (**Supplementary Fig. 5I**).

Having established the identity of these cells, we next asked whether the identified cell type exhibited disease-associated functional programs. Pathway activity analysis across gestational maturity groups and clinical features showed distinct patterns (Fig. 3H): in extreme preterm infants, epithelial-like cells displayed disease-associated changes in apoptosis, inflammatory response, innate-immune epithelial programs, lung development, and oxidative stress. In very preterm infants, EOI and RDS were associated with stronger activation of inflammatory, epithelial barrier, oxidative stress, and cell-cycle programs. In moderate preterm/term infants, pathway activity included inflammatory response, mucociliary differentiation, and epithelial barrier programs.

When focussing on cell–cell communication, networks in preterm infants indicate differential ligand–receptor signaling when comparing differing degrees of immaturity (very and extreme prematurity) indicating a predominant upregulation of signaling in extreme premature in PBMC (top panel) and DPA (bottom panel) and an abrogation of this cell-cell communication with BPD severity, highlighting lung ciliated cells as a central hub in PBMC as well as secretory, basal, and monocyte subsets in DPA.

Orthogonal flow cytometry, single-cell transcriptomics, reference mapping, marker-gene analyses, and pathway-level disease associations identify a circulating respiratory epithelial-like population in neonatal blood associated with developmental immaturity and neonatal lung disease. Rather than representing technical contamination, these cells may reflect epithelial injury, immature barrier function, mucosal shedding, or disruption of the airway–blood interface during early postnatal adaptation. Their enrichment in preterm infants and their disease-associated pathway activity raise the possibility that they provide a blood-accessible window into epithelial stress and lung disease risk during a developmental period when direct sampling from the airway and the blood-gas interface in lung periphery is limited. We treat this finding as a biologically interesting candidate signal that requires further validation in independent cohorts and methods apart from the successful validation in an independent dataset generated on a different technical platform.

### Circulating monocyte signatures are primed by lung epithelial cells via a ciliated-to-myeloid LAMA5–ITGB1 axis in preterm infants with acute lung disease

Having identified compartment-specific myeloid remodeling, we next asked whether airway and circulating myeloid populations were linked through shared transcriptional programs (**Fig. 4A**), revealing substantial transcriptional similarity across compartments.suggesting an across-niche phenotype. In DPA, the “Mono_Recruited” subtype exhibited the strongest correspondence with circulating non-classical monocytes (NCM) and migratory VCAN+ classical monocytes in PBMC. This convergent profile suggests that DPA-recruited monocytes share transcriptional features with (primed) circulating monocytes, consistent with, although not proving active recruitment of myeloid cells from the circulation into the inflamed neonatal airway and/or their later extravasation.

**Figure 4.**
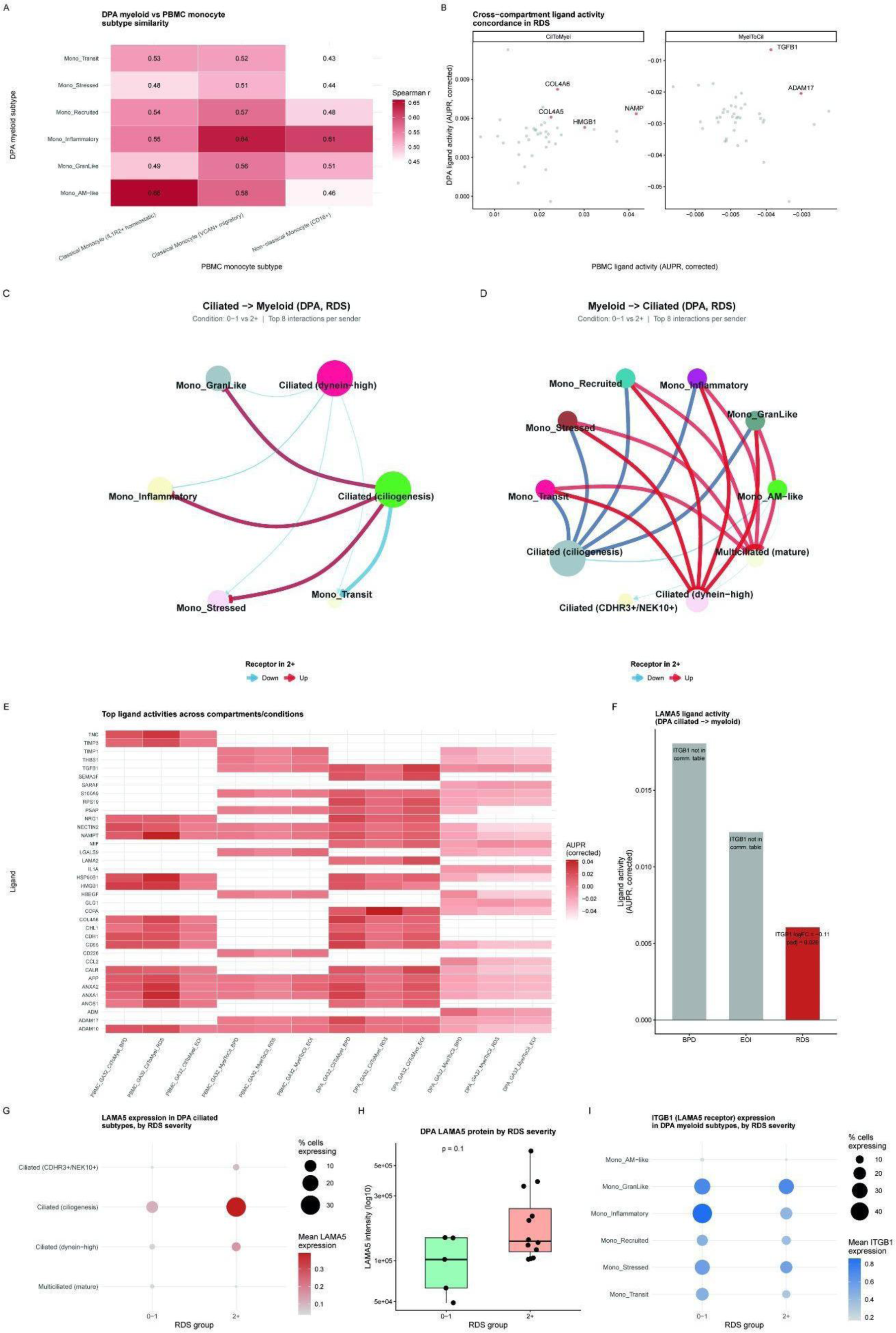
Integrated compartment migration and communication overview links DPA ciliated-myeloid signaling to a concordant, disease-associated LAMA5–ITGB1 axis (GA < 32 weeks). **(A)** Subtype-level similarity (Spearman’s r) between DPA myeloid subtypes and PBMC monocyte subtypes, based on shared transcriptional identity. **(B)** Concordance of NicheNet ligand activity scores (AUPR, corrected) between DPA and PBMC for the ciliated-to-myeloid (CilToMyel, left) and myeloid-to-ciliated (MyelToCil, right) directions in the RDS comparison. Ligands ranking highly in both compartments are highlighted and labeled. (C and D) NicheNet communication networks showing the top 8 predicted ligand–receptor interactions per sending cell type for ciliated→myeloid **(C)** and myeloid→ciliated **(D)** signaling in DPA, comparing RDS 0–1 versus RDS grade >=2 (2+). Edge color indicates the direction of receptor expression change in the RDS grade >=2 (2+) group (blue, downregulated; red, upregulated); edge width reflects interaction strength. **(E)** Heatmap of NicheNet ligand activity scores (AUPR, corrected) for the top-ranked ligands across both compartments (PBMC, DPA), both communication directions (CilToMyel, MyelToCil), and all three clinical comparisons (BPD, RDS, EOI). **(F)** LAMA5 ligand activity score (DPA ciliated→myeloid) across the three clinical comparisons. The RDS bar is highlighted in red, the condition in which the LAMA5–ITGB1 interaction was identified in the NicheNet communication table; annotations show the ITGB1 receptor log2 fold change and adjusted p value for comparisons in which the pair was detected. **(G)** LAMA5 expression in DPA ciliated cell subtypes, stratified by RDS severity. Dot color denotes mean expression; dot size denotes the percentage of cells expressing LAMA5. **(H)** LAMA5 protein abundance in DPA, by RDS severity, quantified by mass spectrometry-based proteomics (six lowest-expressing samples in the RDS 2+ group excluded as technical outliers). p value, two-sided Wilcoxon rank-sum test. **(I)** ITGB1 (the LAMA5 receptor) expression in DPA myeloid cell subtypes, by RDS severity, displayed as in (G). Abbreviations: DPA, deep pharyngeal aspirates; PBMC, peripheral blood mononuclear cell; GA, gestational age; RDS, respiratory distress syndrome; BPD, bronchopulmonary dysplasia; EOI, early-onset infection; AUPR, area under the precision-recall curve; CilToMyel, ciliated-to-myeloid signaling direction; MyelToCil, myeloid-to-ciliated signaling direction; AM, alveolar macrophage; logFC, log2 fold change; padj, Benjamini-Hochberg-adjusted p value.

To investigate how airway epithelial cells might shape these myeloid states, we next examined cross-compartment cell–cell communication. Monocyte subtypes central to early postnatal immune adaptation showed strong incoming and outgoing interaction signals with epithelial populations in DPA (**Fig. 4B,C**). Cell–cell communication analysis with NicheNet revealed predicted signaling between epithelial-like cells and recruited and granulocyte-like monocyte subsets, with directionally consistent patterns across compartments and clinical phenotypes, including BPD and EOI (**Fig. 4D; Fig. S4A,B**). These analyses revealed coordinated epithelial–myeloid communication associated with severe respiratory disease.

Ligand activity analysis (top ligands per comparison; **Fig. 5E**) identified LAMA5 as a candidate driver of ciliated-to-myeloid signaling in preterm infants with respiratory distress syndrome (RDS). Within the predicted LAMA5 signaling network (**Fig. 5F**), receiver monocytes expressed the cognate receptor ITGB1, whose expression was modestly but significantly lower in infants with RDS grade >=2 (2+) than in those with RDS 0–1 (log2FC = −0.11; padj = 0.026). LAMA5 expression in DPA was concentrated in ciliogenesis-program ciliated cells and increased with RDS severity (**Fig. 5G**), and LAMA5 protein in DPA was likewise higher in infants with RDS grade >=2 (2+) than in those with RDS 0–1 (**Fig. 5H**; nominal P = 0.10).

**Figure 5.**
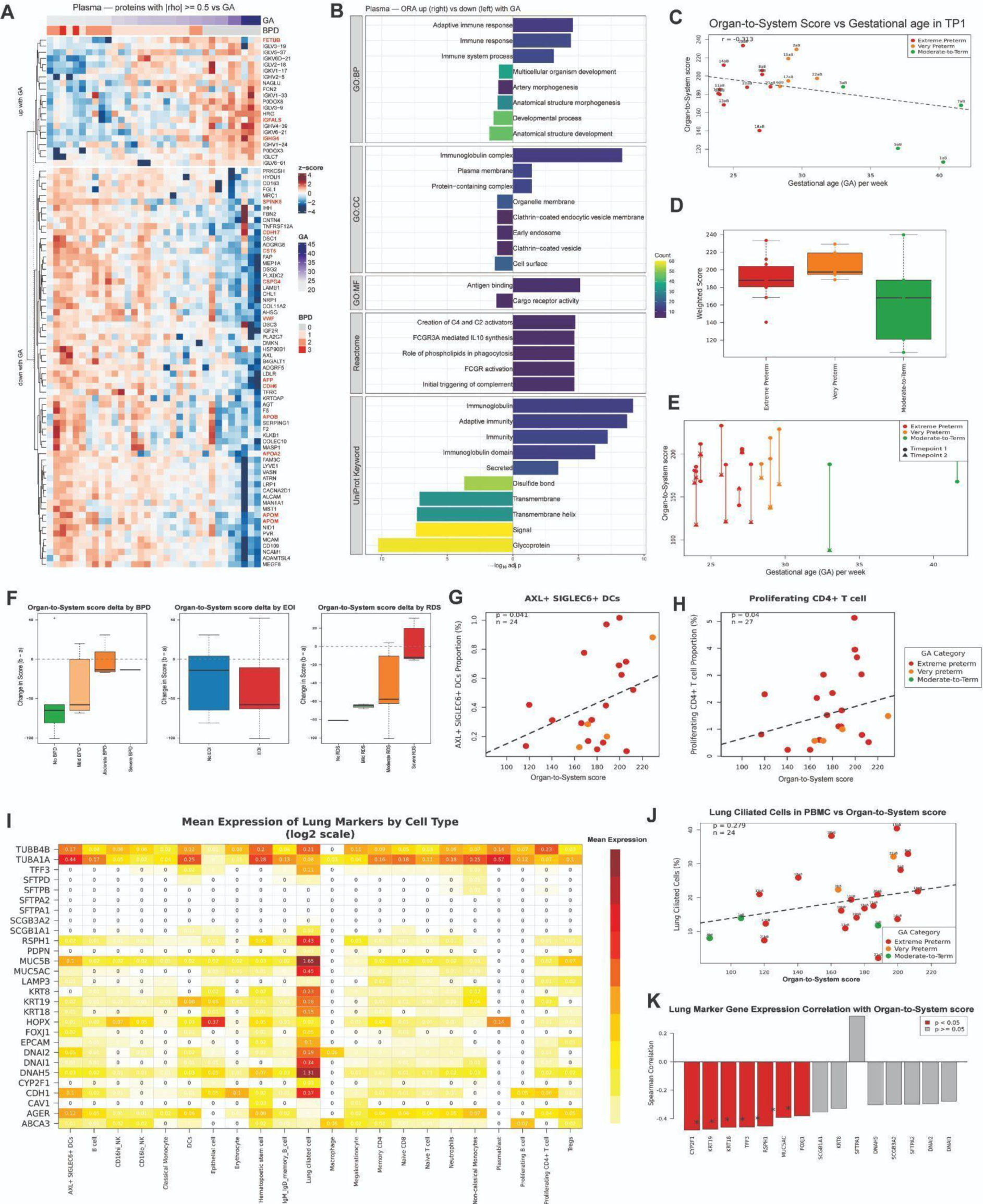
Proteomic and cellular correlates of neonatal organ-to-system score across gestational age and disease severity. **(A)** Heatmap of plasma proteins with |Spearman ρ| ≥ 0.5 versus gestational age (GA), clustered into proteins increasing with GA (top block) and decreasing with GA (bottom block). Columns are samples, annotated by GA (continuous) and BPD grade (0–3) color bars at top; tile color, z-scored protein abundance. **(B)** Over-representation analysis (ORA) of plasma proteins increasing (right-facing bars) versus decreasing (left-facing bars) with GA, across GO Biological Process, GO Cellular Component, GO Molecular Function, Reactome, and UniProt Keyword databases. Bar length, −log10 adjusted p-value; bar color, gene count per term. **(C)** Organ-to-System Score versus gestational age at timepoint 1 (TP1, day of life 1-3), colored by maturity category (Extreme Preterm, Very Preterm, Moderate-to-Term); points labeled by donor ID. Dashed line, linear fit (Spearman r = −0.313). **(D)** Organ-to-System (weighted) Score by maturity category at TP1. Boxplots show median and interquartile range with individual donor values overlaid. **(E)** Organ-to-System Score versus gestational age, showing paired measurements per donor at Timepoint 1 (circle) and Timepoint 2 (triangle), connected by a vertical line; colored by maturity category. (F) Change in Organ-to-System Score between timepoints (Δ = Timepoint 2 − Timepoint 1) stratified by BPD grade (left), EOI status (middle), and RDS grade (right). **(G)** AXL+ SIGLEC6+ DC proportion (%) versus Organ-to-System Score across donors, colored by maturity category; p = 0.041, n = 24. Dashed line, linear fit. **(H)** Proliferating CD4+ T cell proportion (%) versus Organ-to-System Score, as in (G); p = 0.04, n = 27. **(I)** Heatmap of mean lung-marker gene expression (log2 scale) across immune and epithelial cell types from single-cell data. Rows, lung-marker genes (including ciliated markers TUBB4B/TUBA1A/DNAH5/RSPH1/FOXJ1, AT2 markers SFTPA1/SFTPA2/SFTPB/SFTPD/ABCA3, and other epithelial markers); columns, cell types; numbers/tile color, mean expression. **(J)** Lung ciliated cell proportion (%) in PBMC versus Organ-to-System Score, colored by maturity category; p = 0.279, n = 24; points labeled by donor ID. **(K)** Spearman correlation between individual lung-marker gene expression and Organ-to-System Score, ranked by correlation strength. Red bars, p < 0.05; gray bars, p ≥ 0.05. *Abbreviations:*GA, gestational age; TP1, timepoint 1 (day of life 1-3) and 2 (day of life 4-10); BPD, bronchopulmonary dysplasia; EOI, early-onset infection; RDS, respiratory distress syndrome; PBMC, peripheral blood mononuclear cell; DC(s), dendritic cell(s); NK, natural killer cell; ORA, over-representation analysis; GO-BP/CC/MF, Gene Ontology Biological Process/Cellular Component/Molecular Function; Reactome, Reactome pathway database; UniProt, Universal Protein Resource; FCGR3A, Fc gamma receptor IIIa; IL10, interleukin-10; C4/C2, complement component 4/2; ρ, Spearman correlation coefficient; adj.p, adjusted p-value.

Together, transcriptomic similarity, inferred ligand–receptor communication, and orthogonal protein measurements converge on a candidate ciliated-to-myeloid LAMA5–ITGB1 signaling axis associated with RDS severity in preterm infants. Notably, while LAMA5 ligand levels rose concordantly at both the transcript and protein level with RDS severity, expression of its receptor ITGB1 on myeloid cells moved in the opposite direction, a pattern consistent with receptor downregulation following sustained ligand engagement, rather than reflecting a loss of signaling capacity. Although these findings support a biologically plausible model of airway epithelial-to-myeloid communication, functional studies will be required to establish causality.

### DPA-lung cross-compartment NicheNet predicts disease- and maturation-associated intercellular signaling axes

Having identified candidate epithelial–myeloid signaling within the DPA compartment, we next asked whether DPA cell states in disease and prematurity contexts carry predicted signaling potential towards neonatal lung parenchymal cells. To this end, we integrated DPA cell states from our cohort with cell types from the independent neonatal lung single-cell atlas used above for circulating ciliated cell mapping (GSE275938; 67,419 cells^39^), and applied NicheNet in both DPA-to-Lung and Lung-to-DPA directions, stratified by BPD severity and gestational maturity (**Fig. S7**).

In the maturation comparison (extreme preterm vs. moderate+term), the highest-ranked predicted interaction was GDF11-ACVR1B in the Lung-to-DPA direction (AUPR=0.53), consistent with a role for TGF-β superfamily signaling in coordinating alveolar–airway maturation programs across compartments. In the BPD severity comparison and maturation context, CALR– and CEACAM1–HAVCR2 interactions were recurrently predicted in the DPA-to-Lung direction, implicating epithelial ER-stress signaling and immune checkpoint activation as candidate mechanisms by which DPA-derived signals may shape lung myeloid programs in preterm infants with respiratory disease. The bipartite communication networks confirmed Secretory/Basal DPA cells as principal predicted senders and Monocyte/DC Lung cells as principal receivers across analyses. Together, these cross-compartment predictions successfully extend the LAMA5–ITGB1 axis identified above by nominating additional DPA-derived epithelial signals with predicted activity in the neonatal lung and prioritize GDF11, CALR, and CEACAM1 for validation in models of neonatal lung disease.

### Plasma proteomics links gestational maturity, lung injury, and circulating epithelial programs

Having identified coordinated airway–blood immune remodeling, we next asked whether systemic plasma proteins capture this process and provide blood-accessible markers of developmental maturity and lung injury. Mass spectrometry-based plasma proteomics identified a dominant axis of protein abundance variation associated with gestational maturation. Hierarchical clustering segregated donors broadly by BPD grade **(Fig. 5A)**. Restricting analysis to proteins with |r|>0.5 Spearman correlation with GA identified 16 maturation-associated proteins **(Fig. 5B)**, including fetal liver secretory factors (FETUB, IGFALS, IGFBP3), immunoglobulins (IGHG4), and apolipoproteins (APOA2, APOB, APOM, ADIPOQ) as negatively correlated, and proteins of extracellular matrix remodeling (VWF, CDH6, CSPG4) as positively correlated with BPD. Gene ontology enrichment confirmed immunoglobulin complex, antigen binding, and adaptive immune response annotations among GA-positively correlated proteins, with transmembrane and glycoprotein terms predominating among negatively correlated proteins **(Fig. 5C)**. These data establish that the neonatal plasma proteome in very and extremely premature infants primarily encodes a disease-related variation superimposed upon a GA-dominated axis.

To quantify respiratory disease burden in a proteomic framework, we derived a composite weighted Organ-to-System Score from the plasma proteome. For each blood sample, we quantified log2-transformed intensities of a curated panel of lung-cell-type marker proteins spanning ciliated cells (FOXJ1, DNAI1, DNAI2, DNAH5, RSPH1, TUBA1A, TUBB4B), club cells (SCGB1A1, SCGB3A2, CYP2F1), alveolar type 1 cells (AGER, PDPN, CAV1, HOPX), alveolar type 2 cells (SFTPA1, SFTPA2, SFTPB, SFTPC, SFTPD, ABCA3, LAMP3), goblet/mucus cells (MUC5AC, MUC5B, TFF3), and general airway epithelium (EPCAM, CDH1, KRT8, KRT18, KRT19). The score was computed as the weighted sum of detected marker intensities, with ciliated-cell markers weighted 3-fold and alveolar type 2 markers weighted 2-fold relative to the remaining categories, reflecting their greater specificity to distal airway and alveolar epithelium, respectively. This weighting reflects cell-type specificity rather than a statistically derived parameter. The score was calculated independently at each postnatal timepoint and related to gestational age, BPD/RDS severity, and circulating immune-cell composition. The score combines the abundance of selected plasma proteins associated with lung development and injury, weighted by the strength of their univariate association with BPD and RDS severity in our cohort (**Fig. 5C,D**). At the early postnatal timepoint (within the first 72 hours of life), Organ-to-System Score values stratified infants by gestational maturity (**Fig. 5D–E**), with infants born before 28 weeks GA showing the highest and most variable scores, consistent with their greater respiratory morbidity. Paired longitudinal comparison (**Fig. 5F**) showed declining Organ-to-System Score values in most infants between the early and later postnatal timepoints, with the largest reductions in more mature infants — consistent with resolving lung injury in this group.

Correlating PBMC immune-cell proportions with the Organ-to-System Score identified two populations with nominal positive associations: AXL+SIGLEC6+ dendritic cells (**Fig. 5G**; n=24, p=0.041) and proliferating CD4+ T cells (**Fig. 5H**; n=27, p=0.04). These are nominal P-values without multiple-testing correction and should be interpreted as exploratory associations from our single cell cohort.

Interrogating lung-epithelial marker expression across PBMC cell types identified a discrete lung ciliated cell cluster in the circulation (**Fig. 5I**), selectively expressing canonical ciliated markers (EPCAM, FOXJ1, TPPP3, TUBB4B, DNAH5, RSPH1, RSPH4A, PIFO, SNTN, CDC20B, DEUP1) and consistent with the population identified in Figure 3. The proportion of circulating lung ciliated epithelial-like cells correlated with Organ-to-System Score (**Fig. 5J**), suggesting that this circulating population may track with proteomic evidence of lung injury and provides a blood-accessible candidate signal for monitoring lung disease in preterm infants.

### Plasma protein trajectories distinguish acute-to-chronic lung disease and carry exploratory prognostic signals from the earliest timepoint

To dissect the disease-related plasma proteomic signature further, we next asked whether early plasma signatures distinguish infants who progress from acute respiratory disease to chronic lung injury in premature babies (<32 weeks). We defined three donor-level disease trajectories based on RDS and BPD severity: Healthy (RDS 0–1, BPD 0–1; n = 5), Acute-only (RDS grade >=2 (2+), BPD 0–1; n = 7), and Acute-to-chronic (RDS grade >=2 (2+), BPD moderate and severe (2+); n = 5) (**Fig. 6A**). Differential plasma protein analysis identified distinct one-versus-rest signatures characterizing each trajectory in the first week after birth (**Fig. 6B**). We emphasize that the per-group sample sizes are small and the signatures are exploratory and replication in an independent neonatal cohort is required before any prognostic use can be claimed. Focusing on the top 10 proteins per trajectory (**Fig. 6B**), the Acute-to-chronic group showed downregulation of proteins involved in cellular stress response (VNN1, DDI2, RAD23A) and immune maturation (FLII, CXCL16, PIK3IP1) compared with the other two trajectories, suggesting that infants progressing to chronic lung disease exhibit very early alterations in stress-response and immune maturation programs, preceding clinical diagnosis by weeks.

**Figure 6.**
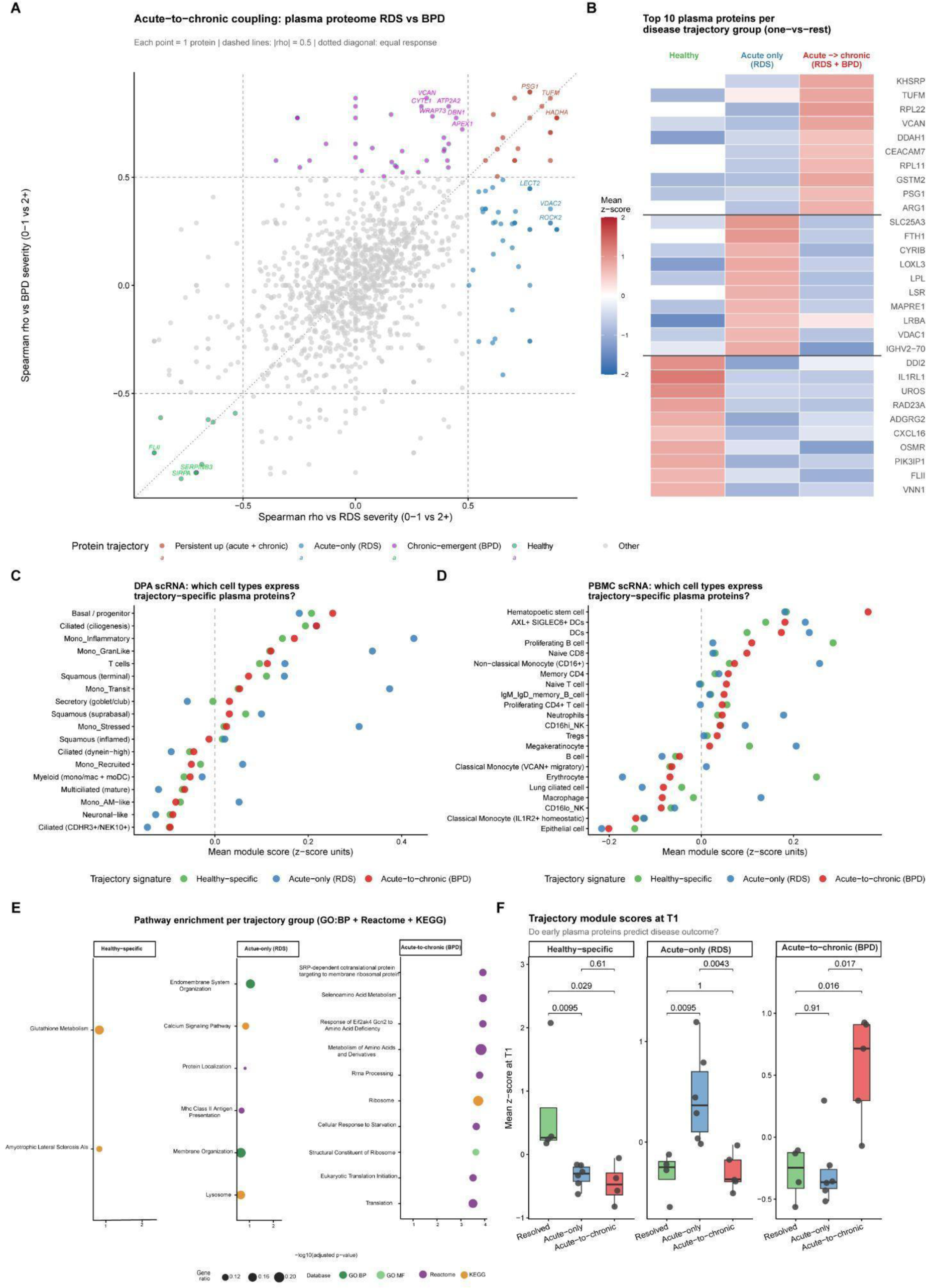
Disease-trajectory groups are defined by donor-level RDS and BPD severity scores for premature baby (<32 weeks): Healthy (RDS 0–1, BPD 0–1; n=5), Acute-only (RDS 2+, BPD 0–1; n=7), and Acute-to-chronic (RDS 2+, BPD 2+; n=5). **(A)** Acute-to-chronic coupling of the plasma proteome. For each plasma protein passing detection/variance filtering (STAR Methods), Spearman’s rho between protein abundance and BPD severity (0–1 vs. 2+) is plotted against Spearman’s rho with RDS severity (0–1 vs. 2+), computed across all plasma samples in both timepoints. Proteins are colored by trajectory module: persistently elevated in both acute RDS and chronic BPD (rho_RDS ≥ 0.5 and rho_BPD ≥ 0.5), RDS-only (rho_RDS ≥ 0.5, rho_BPD < 0.5), BPD-emergent (rho_RDS < 0.5, rho_BPD ≥ 0.5), persistently downregulated (rho_RDS ≤ −0.5 and rho_BPD ≤ −0.5), or other. Dashed lines mark |rho| = 0.5; dotted diagonal indicates equal RDS/BPD association. **(B)** Heatmap of the top 10 plasma proteins most specifically associated with each disease-trajectory group in both timepoints, identified by one-vs-rest Spearman correlation between protein abundance and group membership (rho ≥ 0.5; STAR Methods). Color shows the group-wise mean z-scored abundance across Healthy, Acute-only (RDS), and Acute-to-chronic (RDS+BPD) donors. **(C, D)** Mean module score (average z-scored expression of each trajectory-specific plasma-protein signature from B) across DPA (C) and PBMC (D) scRNA-seq cell types, identifying which airway and immune populations express the transcripts encoding the trajectory-specific plasma proteins for each signature in both timepoints (Healthy-specific, Acute-only/RDS, Acute-to-chronic/BPD). **(E)** Over-representation analysis of the same three one-vs-rest plasma-protein signatures in both timepoints against GO Biological Process, GO Molecular Function, Reactome, and KEGG gene sets (hypergeometric test; clusterProfiler::enricher; BH-adjusted p < 0.25; top 10 terms per group by adjusted p value). **(F)** Trajectory module scores (mean z-scored plasma protein abundance per signature) measured at the earliest sampled timepoint (T1, day of life 1-3), stratified by eventual disease-trajectory outcome. Boxplots show median and interquartile range; points are individual donors. P values are from two-sided Wilcoxon rank-sum tests for each pairwise comparison. Abbreviations used: BPD, bronchopulmonary dysplasia; DPA, distal pulmonary airway; GO, Gene Ontology; KEGG, Kyoto Encyclopedia of Genes and Genomes; PBMC, peripheral blood mononuclear cells; RDS, respiratory distress syndrome; scRNA-seq, single-cell RNA sequencing.

To identify potential cellular sources of these circulating proteins, we mapped trajectory-specific plasma signatures onto the paired airway and blood single-cell atlases. Corresponding genes localized predominantly to ciliated and monocytic cells in DPA (**Fig. 6C**) and adaptive immune populations in PBMC (**Fig. 6D**), providing a cellular framework linking plasma biomarkers to coordinated airway and systemic immune responses. Pathway enrichment of the trajectory-specific protein signatures (**Fig. 6E**) suggested distinct biological programs: cell survival, membrane reorganization, and antigen presentation in the Healthy group; acute innate inflammatory programs in the Acute-only group; and impaired stress and developmental responses in the Acute-to-chronic group.

Finally, we asked whether the trajectory-specific signatures were already detectable at the earliest postnatal timepoint. Module scores calculated from plasma collected within the first 72 hours of life differed across eventual disease trajectories (**Fig. 6F**). Module scores for each trajectory-specific protein signature, computed on the earliest sampled plasma proteome (T1, within the first 72 h of life), differed across eventual disease-trajectory groups (**Fig. 6F**). These findings suggest that early plasma proteomic signatures capture biological programs associated with subsequent respiratory outcomes and nominate candidate prognostic biomarkers for future validation. The analyses extend the airway–blood axis from a descriptive framework of neonatal immune adaptation toward an integrated platform for early risk stratification and biomarker discovery. Detection of the plasma protein trajectory signal at the earliest postnatal timepoint suggests its value as an exploratory prognostic candidate, pending replication in an external cohort.

Taken together, our data support an integrated model in which developmental immaturity propagates across the airway–blood axis to shape early respiratory outcomes (Fig. 7). Lower gestational age was associated with structural and immune immaturity of the airway, comprising immature epithelial barrier, extracellular matrix, ciliary and stress-response programs (Fig. 1, Fig. 3). Within this environment, immature and recruited myeloid populations expanded and acquired compartment-specific activation states spanning recruited, inflammatory, AM-like, stressed/transitional and granulocyte-like phenotypes (Fig. 2). Epithelial–myeloid crosstalk, exemplified by the ciliated cell-derived LAMA5–ITGB1 axis, connected these programs across the epithelial barrier and was concordant at the transcript and protein level with RDS severity (Fig. 4). Loss of airway– blood compartmentalization was reflected systemically by circulating respiratory epithelial-like cells co-expressing EPCAM, FOXJ1, TPPP3, RSPH1 and DNAH5, and by lung-associated remodeling of the plasma proteome (Fig. 3, Fig. 5). The magnitude and persistence of this dysregulation tracked with clinical course, separating healthy, acute-only (RDS) and acute-to-chronic (RDS + BPD) trajectories and yielding blood-accessible candidate readouts detectable within the first 72 h of life (Fig. 6). This model synthesizes associative findings from a single cohort and is intended as a hypothesis-generating framework rather than an established causal sequence.

**Figure 7.**
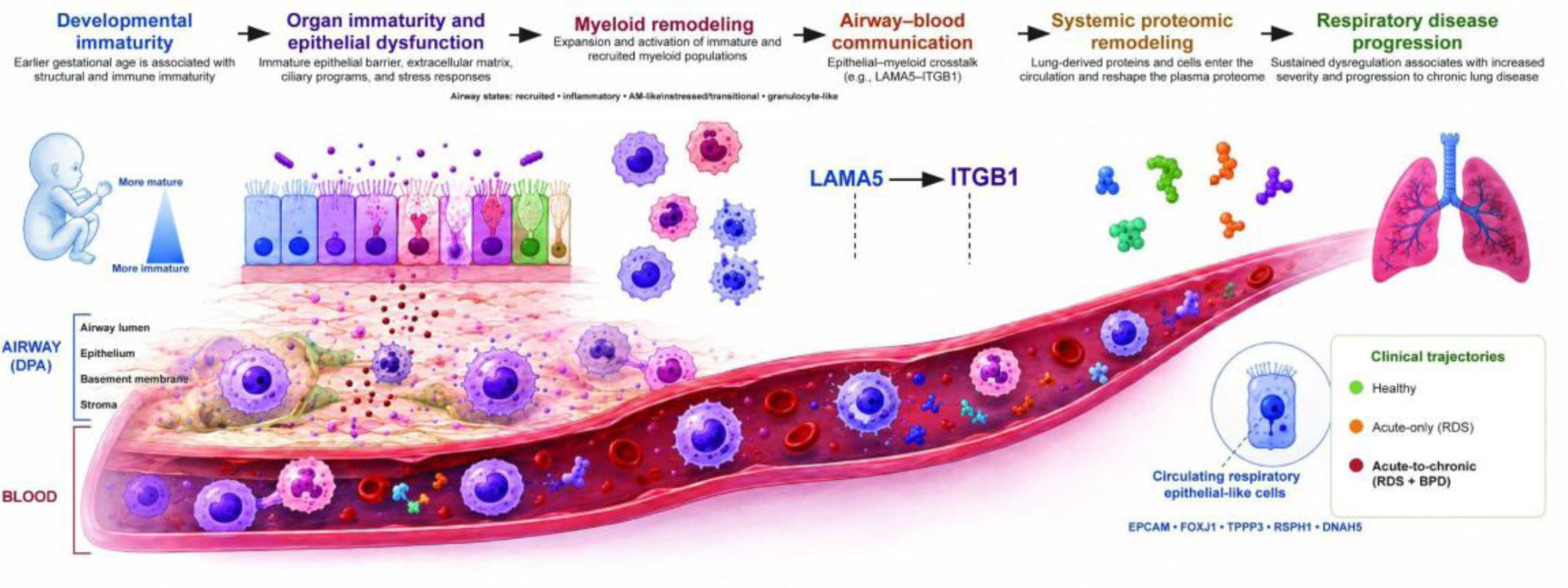
Integrated model of the neonatal airway–blood axis linking gestational immaturity to respiratory disease trajectory. Schematic summary of the study, shown across the airway (DPA, top) and blood (bottom) compartments. Left to right: (i) Developmental immaturity, earlier gestational age is associated with structural and immune immaturity; (ii) Organ immaturity and epithelial dysfunction, immature epithelial barrier, extracellular matrix, ciliary and stress-response programs across the airway lumen, epithelium, basement membrane and stroma; (iii) Myeloid remodeling, expansion and activation of immature and recruited myeloid populations, with airway states spanning recruited, inflammatory, AM-like, stressed/transitional and granulocyte-like phenotypes; (iv) Airway–blood communication, epithelial–myeloid crosstalk, including the ciliated cell-derived LAMA5–ITGB1 axis; (v) Systemic proteomic remodeling, lung-derived proteins and circulating respiratory epithelial-like cells (EPCAM, FOXJ1, TPPP3, RSPH1, DNAH5) enter the circulation and reshape the plasma proteome; (vi) Respiratory disease progression, sustained dysregulation associates with greater severity and progression to chronic lung disease, distinguishing healthy, acute-only (RDS) and acute-to-chronic (RDS + BPD) clinical trajectories. Created with BioRender.com. Abbreviations: DPA, deep pharyngeal aspirate; AM, alveolar macrophage; ECM, extracellular matrix; LAMA5, laminin subunit alpha 5; ITGB1, integrin beta 1; EPCAM, epithelial cell adhesion molecule; FOXJ1, forkhead box J1; TPPP3, tubulin polymerization-promoting protein family member 3; RSPH1, radial spoke head 1 homolog; DNAH5, dynein axonemal heavy chain 5; RDS, respiratory distress syndrome; BPD, bronchopulmonary dysplasia.

## Discussion

The continuous crosstalk between host and environment is shaped in the “window of opportunity” early after birth^40^ and particularly important yet challenging to study in preterm infants in which early immune dysregulation defines long-term outcome^41^. Our study establishes the neonatal airway–blood axis as an integrated framework for understanding early postnatal immune adaptation. By combining paired airway and blood single-cell transcriptomics with plasma proteomics, we show that developmental immaturity remodels coordinated, compartment-specific immune programs linked to respiratory disease.

**Three conceptual advances** emerge as the main contributions of this pilot study. ***First,*** gestational maturity is associated with compartment-specific patterns of immune composition and pathway activation in airway and blood, with extreme preterm infants showing the most divergent profiles. ***Second,*** we identify and validate a circulating respiratory epithelial-like population in neonatal blood that is enriched in preterm infants and associated with disease-related pathway activity. ***Third,*** plasma proteomics surfaces an Organ-to-System Score and a set of plasma protein trajectories that track gestational maturity and lung disease severity. The discussion addresses each of these points in turn before outlining the limitations of the study and pointing to priority directions for replication and functional follow-up. The graphical abstract (**Fig. 7**) conceptualizes the scientific advancements in a schematic fashion.

### Developmental immaturity remodels coordinated immune adaptation across the airway–blood axis

Our integrated dataset combined with the fine-granulated segregation into gestational age groups allowed us to identify distinct immune phenotypes during early postnatal life as well as to characterize distinct trajectorial changes upon injury and disease development. In extreme prematurity, classical and non-classical monocytes are abundant and exhibit activation programs (IL-1, TNF–NF-κB, interferon-associated, glycolytic, oxidative-stress) that differ from more mature infants. In parallel, the lymphocyte compartment shows progressive contraction and shifts in T- and NK-cell composition with decreasing gestational age, consistent with delayed adaptive immune development. These patterns are consistent with — though not definitive proof of — the heterogeneity of prematurity as an immune state and underline the need for gestational age-resolved analyses^1,11,12^. In comparison with prior neonatal immune atlases, our paired airway–blood pilot extends conceptual understanding by providing a directly matched sample-level view of compartment-specific immaturity in preterm infants.

The compartment-specific patterns suggest that disease development unfolds along distinct programs in airway and blood, and that gestational age modulates the relative balance of these programs. Although a causal role for any specific program cannot be established from these pilot data, the study’s delineation of important signals directs future studies. Our findings confirm important studies in the field demonstrating GA as a major driver of a perinatal IL-1B/TNF-centered immune response in placental immune cells and the presence of classical monocytes as well as specific T cell subsets^11,12,1,13,14,42^. The lack of an integrated view through paired sampling, however, significantly hindered these studies to identify important cross-talk mechanisms.

Examining the relationship between local airway and systemic blood compartments, we find that myeloid and epithelial cell programs differ between DPA and PBMC regarding cellular composition and pathway activity depending on immaturity. DPA recruited and granulocyte-like monocyte subsets share transcriptional features with non-classical and migratory classical monocytes in blood, suggesting a connected compartment in which primed circulating monocytes may be mobilized to the airway. The candidate LAMA5–ITGB1 axis (Fig. 4) provides a hypothesis for how airway epithelial signals could shape this mobilization, as LAMA5 is known to alter β1 integrin signaling through the non-canonical kinase PYK2 and the skeletal enriched SRC kinase, FYN, impacting on cytoskeleton and WNT signaling^43^.

Although studies identified proteomic signatures^42^, their integration in cross talk mechanism is hindered by the use of targeted approaches as well as the lack of integrated (paired) datasets that allowed us to identify two airway-specific signaling axes (LAMA5-ITGB1, CXCL5-GRM7(data not shown)) and the presence of a circulating epithelial cell.

Our study conceptualizes prematurity as a paraimmune state (^44–48^), in which environmental injuries perpetuate tissue injury and inflammation as well as morbidity development itself tip the immune disturbance towards disease.

### Myeloid and epithelial programs coordinate airway–blood communication

The convergent gestational age and disease associations of classical monocyte abundance, non-classical monocyte expansion, and DPA-recruited monocyte states in this pilot are consistent with prior reports of early myeloid-skewed responses in BPD ^4,9,10^. Data published reflect on the network activation we demonstrated^42^ and highlight IFN-γ, CXCL9, CXCL10 signaling while providing a basis to explain a progressive Th17 polarization (CD161+CD4+CCR6+CCR4+) and neutrophilia that evolves over months into disease development, directly mapping our acute-to-chronic trajectory.

Central to the early postnatal immune responses especially in preterms, the persistent remodeling of myeloid cells likely reflects the effects of environmental hits including oxygen, microbial signals, and mechanical forces, plausibly contributing to tissue injury and sustained local inflammation.

Our data add a paired-compartment view in which airway-recruited monocytes transcriptionally resemble specific circulating monocyte states, hinting at shared developmental or activation programs across compartments.

The identification of a circulating respiratory epithelial-like population in neonatal blood is, to our knowledge, the first explicit description of this signal in a paired airway–blood single-cell dataset of this gestational range. Immune-focused workflows that sort or computationally remove non-immune events would typically have discarded these cells. Their enrichment in preterm infants, their disease-associated pathway activity, and the convergent FACS validation argue for a biologically structured population rather than a technical artifact. We considered, and partially addressed, several alternative explanations (doublets, ambient RNA, sample-handling carry-over). However, orthogonal validation of circulating epithelial/ciliated cells in preterm blood in publicly available blood scRNA-seq dataset from Das et al.^11^ generated on a different platform (GSE236099, 10x CITE-seq, 22,775 PBMCs) in 3 very preterm infants <32 weeks GA by applying the demonstrated combination of 14 ciliated/epithelial markers strongly supports our findings through successful reproduction of the same biological signal. To further anchor the transcriptional identity of these circulating cells to a defined lung cell type, we performed reference-based integration of the PBMC-derived ciliated population onto a published neonatal lung single-cell atlas^39^ (GSE275938; 67,419 cells) using scArches query mapping. The PBMC ciliated cells co-localised almost exclusively with the Shirazi ciliated epithelial cluster, exhibiting a 20.8-fold KNN enrichment with 58.5% of nearest-neighbour contacts mapping to this single cluster. This degree of transcriptional proximity, further supported by the specific expression of the full ciliated marker programme (FOXJ1, DNAH5, TPPP3, PIFO, CDC20B and others) exclusively within this cluster (Supplementary Fig. 5), strongly suggests that the circulating cells originate from, or are transcriptionally very similar to ciliated airway epithelium of the neonatal lung. The route of entry into the circulation, passive shedding across an immature epithelial barrier, active mobilization, or trapping during pulmonary transit, remains to be defined. Further independent validation in cohorts with methylation-based tissue-of-origin profiling or in situ imaging will be required to firmly establish tissue source and clinical relevance of the signal.

Epithelial–immune crosstalk in the neonatal airway and circulation is conceptually attractive but remains incompletely characterized at the molecular level. In our data, candidate ligand–receptor interactions between epithelial-like cells (in airway and blood) and monocyte subsets converge on the LAMA5–ITGB1 axis in preterm infants with lung disease. This convergence supports a working hypothesis for a ciliated-to-myeloid signaling axis but does not establish a causal role; functional validation engaging receptor blockade and ex vivo stimulation as well as confirmation are required. We can thereby significantly add to studies that proposed that preterm immune cells are predominantly “receiving signals” rather than sending, indicating a passive/dysregulated communication network^14^.

Cross-compartment NicheNet analysis integrating DPA cell states with an independent neonatal lung atlas ^39^ further enriched the candidate signaling axes. GDF11 was the top-ranked predicted ligand in the maturation comparison (Lung-to-DPA), signaling via ACVR1B/ACVR2A on DPA epithelial cells. Given the essential role of activin receptor signaling in alveolarization^49^, selective GDF11 prediction in extreme preterms suggests disruption of a physiological lung-to-airway maturation pathway. CEACAM1 also emerged as a leading DPA-to-Lung ligand in the maturation comparison, with HAVCR2 (Tim-3) as the predicted receptor, suggesting a potential role for DPA epithelial cells in suppressing myeloid activation through Tim-3 signaling, complementing the LAMA5–ITGB1 axis. In the BPD severity comparison, C3 was the highest-ranked DPA-to-Lung ligand, predicted to signal through CR1, CD46, and ITGAX on lung myeloid cells, consistent with the contribution of complement activation to BPD pathogenesis ^50^. Conversely, OSM was the top Lung-to-DPA ligand, acting via OSMR/IL6ST (gp130), in agreement with its established pro-inflammatory and pro-fibrotic functions^51^. These predicted interactions require validation in paired DPA–BAL samples and functional models.

### Plasma captures a systemic readout of airway biology and identifies candidate biomarkers of disease progression

Plasma proteomics in this pilot surfaces a coordinated set of gestational-age-associated proteins, including fetal-maturation markers (FETUB, IGFALS, IGFBP3), lipoproteins (APOA2, APOB, APOM, ADIPOQ), and tissue-remodeling proteins (VWF, CDH6, CSPG4). To summarize the collective signal of these and related lung-injury-associated proteins, we computed an Organ-to-System Score — a weighted composite of protein abundances, with weights derived from their association with gestational maturity and lung injury severity within this discovery cohort. The score stratifies infants by gestational maturity and chronic lung disease severity in the same samples from which it was derived, thereby functioning as a descriptive, exploratory index rather than a validated prognostic tool.

Chronic lung disease, clinically only diagnosed weeks to months after birth, with its significant impact on long-term outcome^52^ has to be scaled on a developmental trajectory whose initial conditions are set before birth by immaturity of immune and barrier compartments, and whose path is further shaped by postnatal insults including infection and clinical care. The acute-to-chronic plasma protein signature identified here, detectable at the earliest postnatal timepoint, is consistent with this trajectory view and is an exploratory prognostic candidate pending independent replication. The highlighted, gestational age dependent disbalanced of immune response through e.g. STAT-signaling reflects on previous studies referring to their clinical importance ^53,54^. Comparison with prior plasma proteomic studies of BPD trajectories and prediction models validates the crucial role of myeloids ^55–58^ while expanding current disease understanding and biomarker searches. Our signatures suggest that neonatal lung disease is not associated with a single myeloid activation state, but instead reflects compartment-specific remodeling of circulating and airway-localized myeloid programs in close relation to the local epithelial compartment, thereby providing a novel view on studies showing innate immune cell presence and an imbalanced adaptive immune response in infants with BPD^59,60^. Future therapeutic approaches will benefit from the dataset-derived information on timing and potency to guide e.g. replacement of multipotent cells or their supernatants in ongoing clinical trials^61–63^.

If validated in larger cohorts, the Organ-to-System Score and the Acute-to-chronic plasma protein trajectory could provide blood-accessible, gestational age-specific biomarker candidates of lung disease risk in preterm infants, complementing clinical risk scores. Single proteins point to known factors of fetal growth (IGFBP3, ADIPOQ) ^64,65^, whereas others might serve as much needed indicators of disease endotypes (VWF, CSPG4) ^66,67^. The circulating respiratory epithelial-like population is a distinct candidate signal that may capture epithelial barrier perturbation and can provide a systemic readout of the detrimental environmental signal obtained in the respective compartment. Future studies will need to ascertain independent validation, a defined evaluation framework, and explicit comparison to existing clinical predictors before translational use for each of the candidates. The data and code released with this study aim to enable such validation (Zenodo repository:XX, CELLxGene (<u>link</u>).

#### Limitations and future directions

Limitations include the cohort size, reflecting the difficulty of collecting paired airway, blood, and plasma samples in the first week of life from preterm infants of a broad gestational age range. We therefore interpreted all stratified comparisons as exploratory and hypothesis-generating. The differential abundance comparison of CXCL5 protein in DPA between RDS severity groups is exploratory as it was based on a limited patient number after explicit outlier exclusion. The Organ-to-System Score is derived and evaluated on the identical data, therefore framed as a candidate composite biomarker requiring independent validation. The circulating respiratory epithelial-like population, supported by orthogonal FACS evidence and validated in an independent dataset, has not yet been further validated by methylation-based tissue-of-origin profiling or in situ imaging. Likewise, the route of entry into the circulation remains unresolved. Multiple sample-handling and pre-analytic factors — including DPA sampling depth and timing and plasma processing, as well as FACS panel differences across days could not be fully controlled in a pilot setting. All plasma proteomic, scRNA-seq, and FACS analyses are correlative; no causal mechanism (e.g., for the LAMA5–ITGB1 axis) was demonstrated. Finally several methodological details require clarification or additional analyses as the cohort with limited sample number and volume was recruited at a single site with external validation in independent cohorts with harmonized sampling planned before any prognostic, mechanistic, or translational implication.

*In summary,* this paired airway–blood scRNA-seq and plasma proteomic study in 19 preterm and term infants provides a multi-layered view of immune and epithelial adaptation in the first week of life (see **Fig. 7**). The three main observations — gestational age- and compartment-specific immune patterning, a circulating respiratory epithelial-like population enriched in preterm infants, and a plasma proteomic Organ-to-System Score and Acute-to-chronic trajectory signature — suggest that neonatal lung disease can be viewed as a developmental trajectory shaped by gestational maturity, environmental exposures, and compartment-specific immune programs.

Each of these observations is offered as a hypothesis-generating candidate signal requiring independent validation rather than as an established mechanism or biomarker.

The early myeloid/macrophage dominance and comparatively reduced presence of mature adaptive lymphocytes as gestational age decreases has been described, next to the tissue microenvironment shaping the immune cell phenotype and pays tribute to the fact that non-immune cells are less well captured during dissociation or are discarded in immune-focused workflows. Consistent with this, we demonstrate circulating epithelial-like cells in the blood in addition to immune cells^1,13^.

Next to our validation steps, future studies need to confirm the findings in larger, independent neonatal cohorts with harmonized longitudinal sampling across systemic and airway compartments. Priority directions include: (i) external replication of the Organ-to-System Score and the Acute-to-chronic plasma protein trajectory in cohorts with full long-term morbidity ascertainment; (ii) orthogonal validation of the circulating respiratory epithelial-like population by methylation-based tissue-of-origin profiling and in situ imaging; (iii) functional testing of the candidate LAMA5-ITGB1 ciliated-to-myeloid axis (e.g., receptor blockade, *ex vivo* monocyte stimulation, neonatal animal models); and (iv) integration with clinical predictors and granulated environmental exposure metrics (e.g. mechanical ventilation, oxygen supplementation) to test incremental prognostic value over existing scores.

## Data availability

Single-cell RNA-seq data will be deposited at GEO and is available for interactive exploration at CELLxGENE (<u>link</u>). Plasma proteomic raw and processed data will be deposited at PRIDE/ProteomeXchange. Analysis code is available at Zenado (<u>10.5281/zenodo.21137413</u>). Cohort-level metadata is available from the corresponding author upon reasonable request, subject to appropriate data-sharing agreements.

## Contributions

Conceptualization, HBS, AH; Investigation: AH, CN, ZW, SK, AWF, KF; Methodology: ZW, SK, HBS, KL; Data Curation: SK, KL; Writing - Original Draft: SK, AH, KMB; Writing-Review & Editing: SK, AH, HBS, KMB; Funding Acquisition: AH, HBS, KMB; Resources: HBS, AH; Supervision: AH, HBS, KMB.

## Acknowledgments

We thank the patients and their families for the significant contribution to the study. AH, HBS, CN and SK received financial support for this work from the Deutsche Forschungsgemeinschaft (DFG, German Research Foundation) – TRR 359 – Project No. 491676693 and the Chan Zuckerberg Initiative (2021-237918). AH received support through the Research Training Group ‘Targets in Toxicology’ (GRK2338, German Research Organisation (DFG)) and the German Center for Lung Research (DZL). AH, AH, KL, MW and EM received support for their work from the Bundesministerium für Forschung, Technologie und Raumfahrt (BMFTR) - INGVER-Project (No. 01KX2419). AH, AH, EM, KL received support from the Profilinitiative Early Development of Carl von Ossietzky Universität Oldenburg (No. PRI 2024-01 ZfE). KMB discloses support for the research of this work from the Chan Zuckerberg Initiative (2021-237918), National Institutes of Health–National Cancer Institute (NIH– NCI) Cancer Center Support Grant (#P30CA016059), the VCU Department of Oral and Molecular Craniofacial Biology, Philips Institute for Oral Health Research (start-up funds), and Massey Cancer Center Harrison Scholars Award.

---

**Supplementary Table S1:**
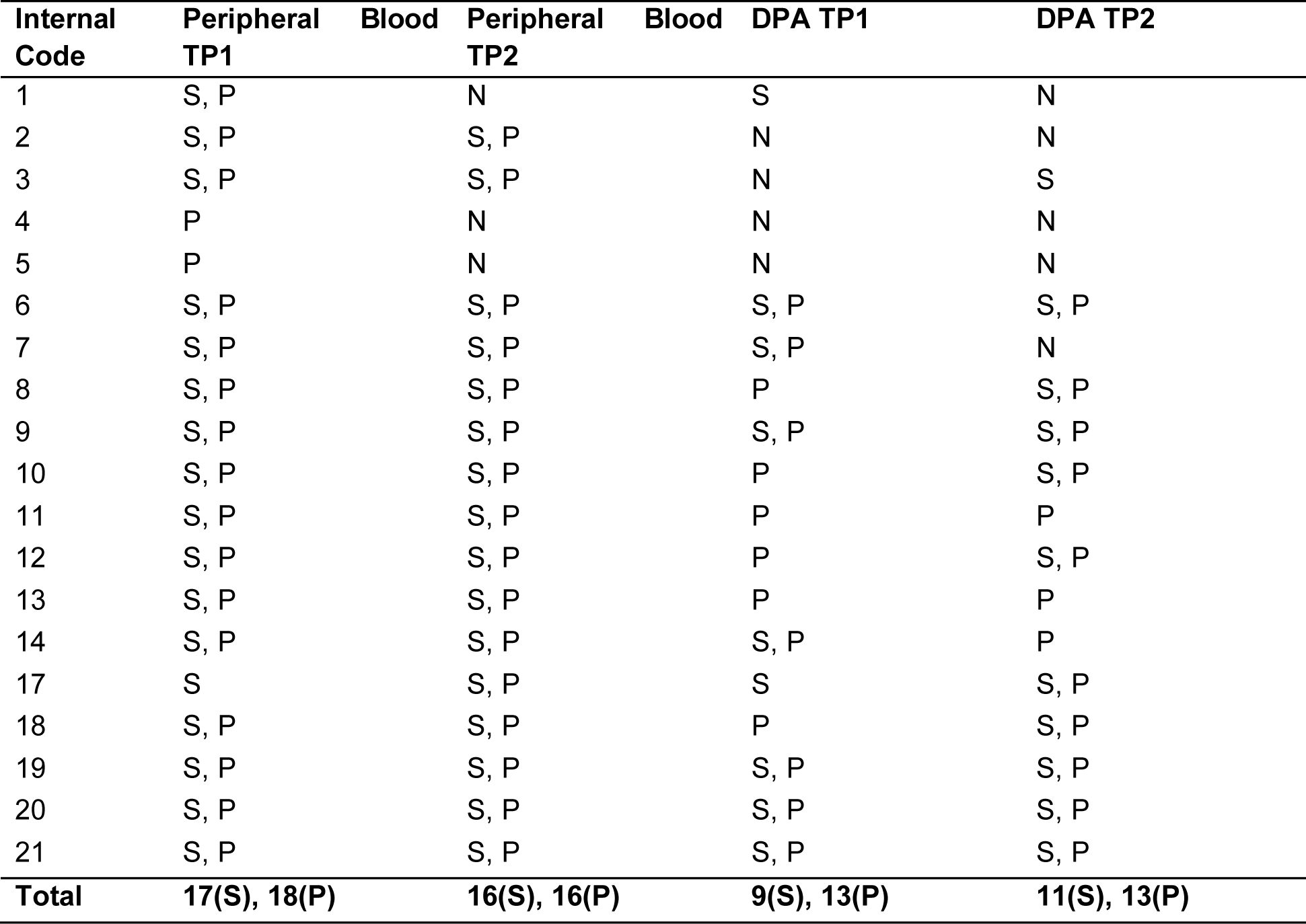
Distribution of Paired and Singleton Samples. Distribution of paired and singleton samples, 19 donors in total; DPA, Deep Pharyngeal Aspirate; N, no sample; P, included in proteomics; S, included in scRNA transcriptomics; TP1 and TP2, sampling timepoint day 1-3 of life (1) and day 4-10 of life (2)

**Supplementary Figure 1.**
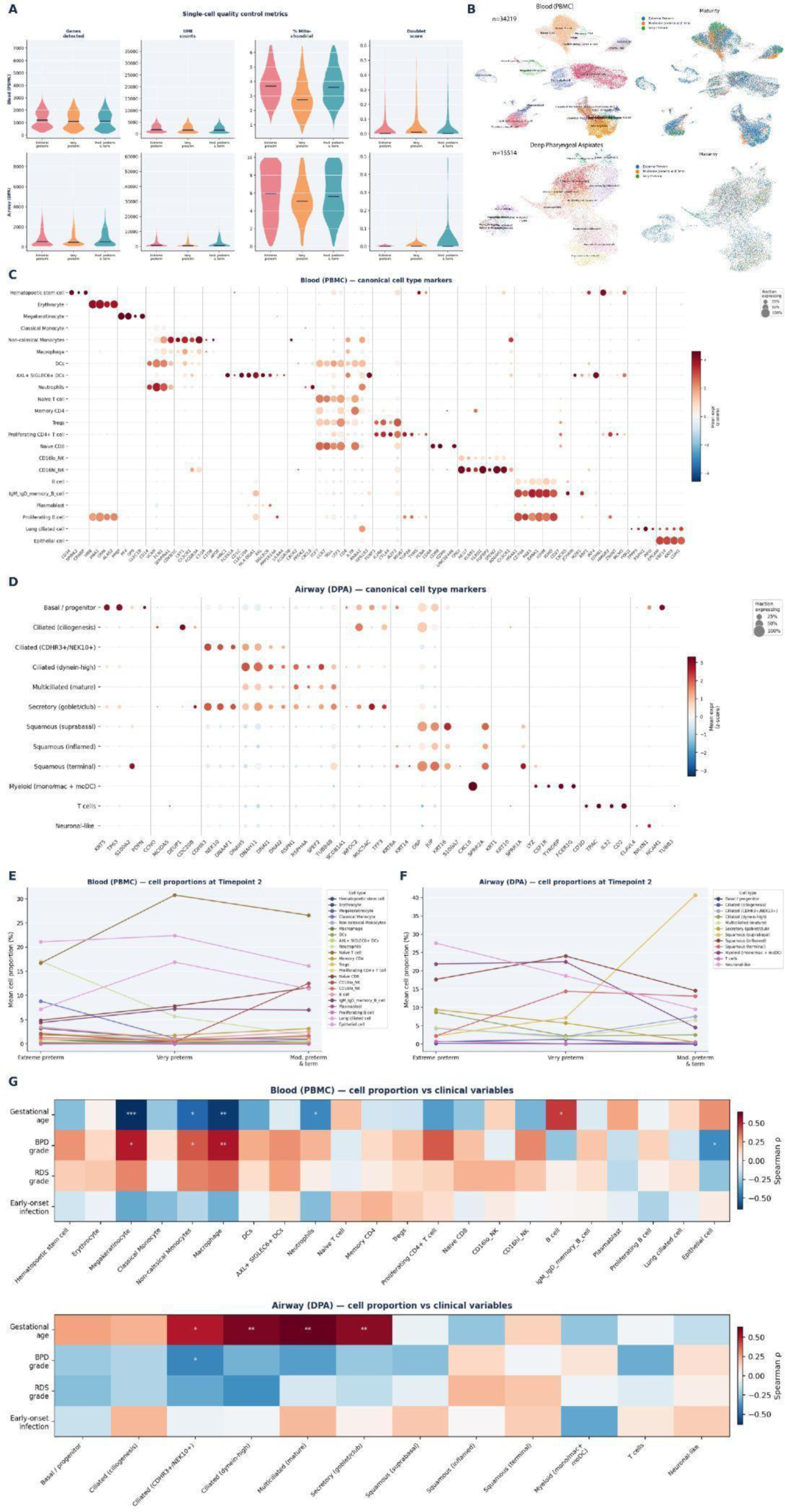
Single-cell quality control and cross-compartment cell-type annotation in blood and airway samples. (A) Single-cell quality-control metrics (genes detected, total counts, percent mitochondrial reads, doublet score) per maturity group for blood (PBMC) and airway (DPA) datasets. (B) UMAP embeddings of blood (PBMC) and airway (DPA) cells, colored by maturity/severity group and disease status. (C) Dot plot of canonical cell-type marker gene expression across blood (PBMC) cell-type clusters; dot size, fraction of cells expressing; color, mean expression (z-scored). (D) Dot plot of canonical cell-type marker gene expression across airway (DPA) cell-type clusters, as in (C). (E) Mean cell-type proportion (%) in blood (PBMC) across maturity groups at Timepoint 2. (F) Mean cell-type proportion (%) in airway (DPA) across maturity groups at Timepoint 2, as in (E). (G) Heatmap of Spearman correlation (ρ) between blood (PBMC) cell-type proportions and clinical variables (gestational age, BPD grade, RDS grade, early-onset infection); *p<0.05, **p<0.01. (H) Heatmap of Spearman correlation (ρ) between airway (DPA) cell-type proportions and clinical variables, as in (G).

**Supplementary Figure 2.**
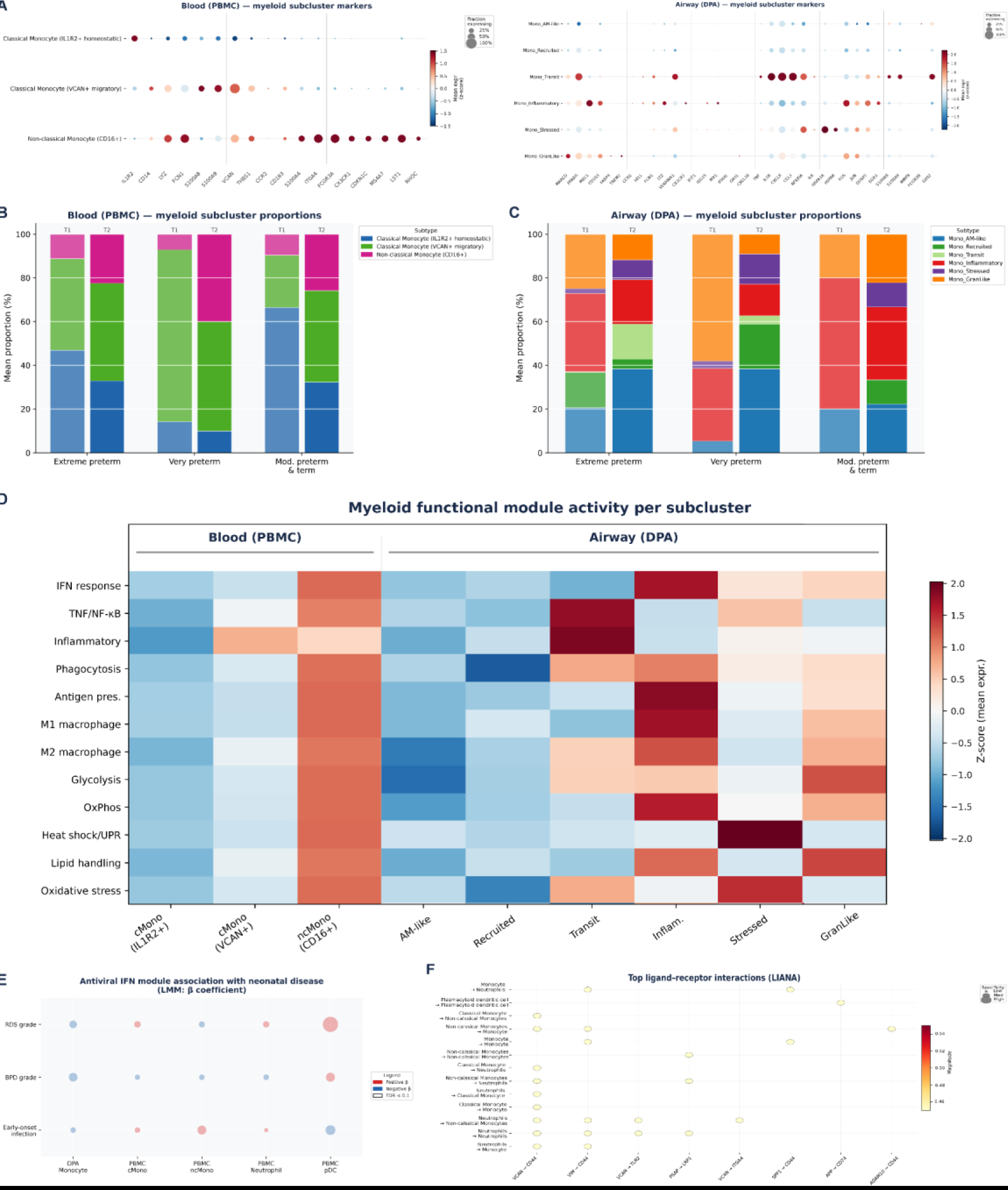
Myeloid/monocyte subclustering and functional module activity in blood and airway compartments. (A) Dot plots of subcluster-defining marker genes for blood (PBMC) classical/non-classical monocyte subclusters (left) and airway (DPA) myeloid subclusters (right). (B) Mean myeloid subcluster proportion (%) in blood (PBMC) across maturity groups. (C) Mean myeloid subcluster proportion (%) in airway (DPA) across maturity groups, as in (B). (D) Heatmap of myeloid functional module activity (z-scored mean module expression) per subcluster in blood (PBMC, left) and airway (DPA, right), covering interferon response, TNF/NF-κB signaling, inflammatory, phagocytosis, antigen presentation, M1/M2 macrophage polarization, glycolysis, oxidative phosphorylation, heat-shock/unfolded protein response, lipid handling, and oxidative stress programs. (E) Association between antiviral interferon module score and neonatal disease severity; dot size/color represents the linear mixed-model β-coefficient and significance. (F) Top ligand-receptor interactions identified by LIANA across myeloid subtypes/conditions; dot size, interaction specificity; color, interaction score

**Supplementary Figure 3.**
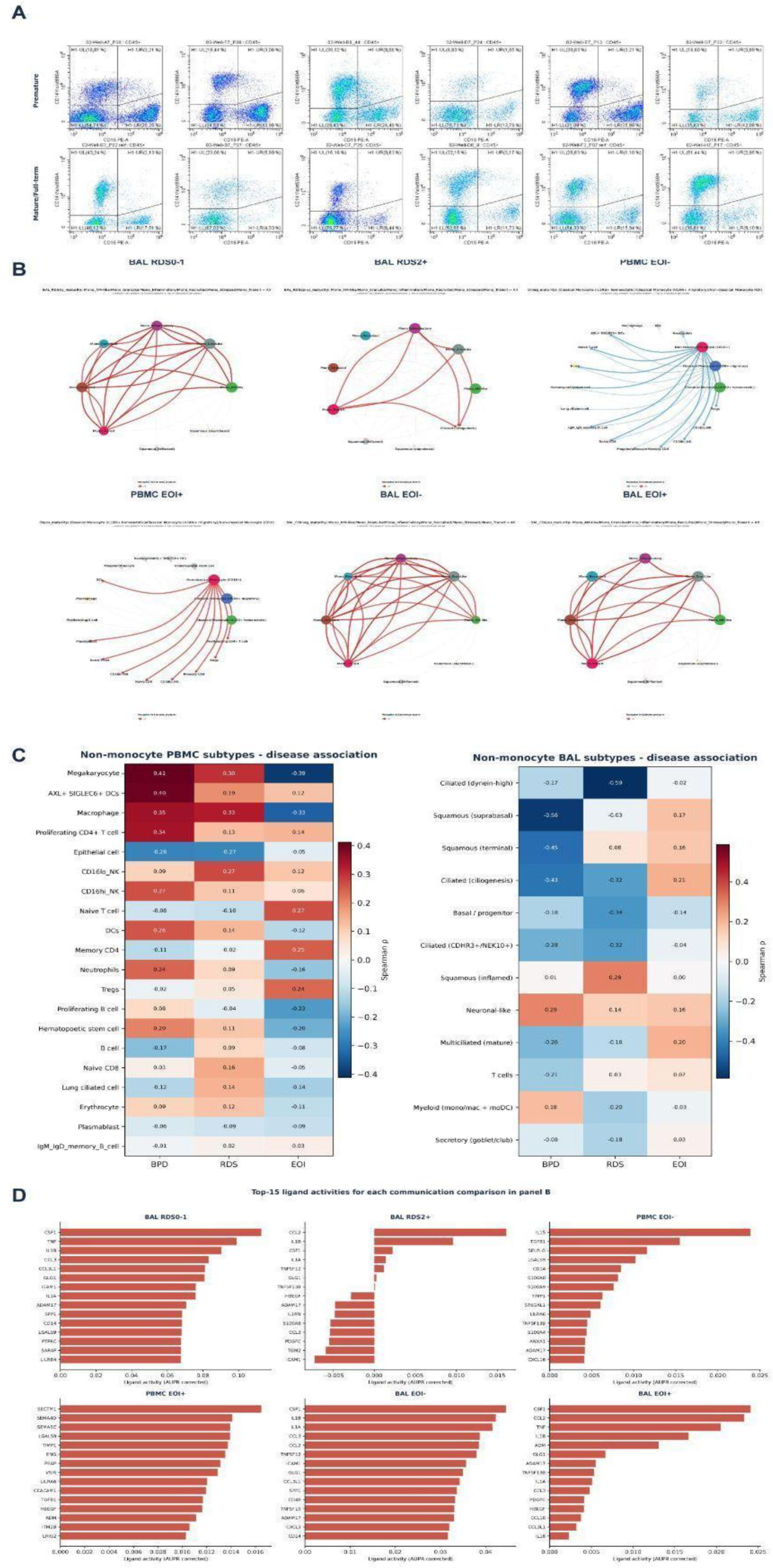
Flow-cytometric validation of myeloid populations and disease-associated ligand–receptor signaling. (A) [unlettered in source] Representative CD14/CD16 flow-cytometry plots of classical/non-classical monocyte gating across individual premature (top row) and mature/full-term (bottom row) infant samples. (B) Circle plots of predicted cell–cell communication networks for BAL RDS0-1, BAL RDS2+, PBMC EOI-, PBMC EOI+, BAL EOI-, and BAL EOI+ groups. Edge color/width denotes interaction strength and direction between cell-type nodes. (C) Heatmaps of Spearman correlation (ρ) between non-monocyte cell-subtype proportions and disease variables (BPD, RDS, EOI) in PBMC (left) and BAL (right) compartments. (D) Bar plots of the top 15 ligand activities (AUPR, corrected) for each communication comparison shown in (B): BAL RDS0-1, BAL RDS2+, PBMC EOI-, PBMC EOI+, BAL EOI-, and BAL EOI+.

**Supplementary Figure 4.**
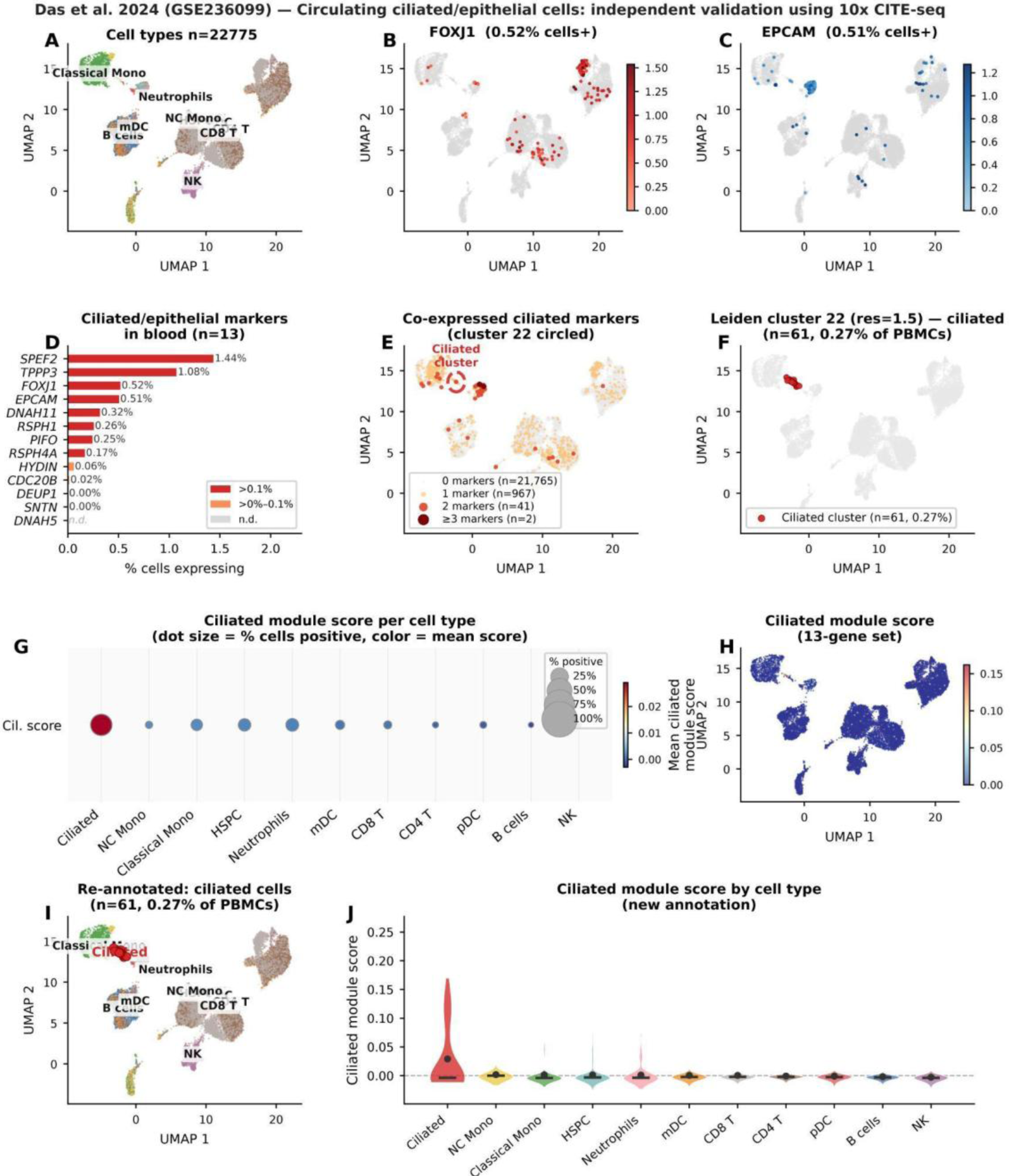
Independent validation of circulating ciliated/epithelial cells in preterm neonatal blood using publicly available 10x CITE-seq data (Das et al., 2024; GSE236099). Re-analysis of 22,775 PBMCs from 3 very preterm infants (<32 weeks GA) profiled by 10x CITE-seq on a platform independent of our cohort. (A) UMAP of all 22,775 PBMCs coloured by annotated cell type, identifying major immune populations including classical monocytes, non-classical monocytes (NC Mono), neutrophils, myeloid dendritic cells (mDC), CD8 T, CD4 T, B cells, and NK cells. (B–C) Expression of ciliated epithelial markers *FOXJ1* (B) and *EPCAM* (C) overlaid on the UMAP. Each gene is detected in approximately 0.5% of PBMCs, with expression concentrated in a discrete cluster located at the top of the UMAP. (D) Percentage of PBMCs expressing each of 13 ciliated/epithelial marker genes. *SPEF2* (1.44%) and *TPPP3* (1.08%) are the most broadly detected, followed by *FOXJ1* (0.52%) and *EPCAM* (0.51%). Bars are coloured by expression prevalence (dark red: >0.1%; light orange: >0%–0.1%). (E) Co-expression map of ciliated markers. Cells are coloured by the number of ciliated marker genes co-expressed simultaneously: 0 markers (n=21,765), 1 marker (n=967), 2 markers (n=41), or ≥3 markers (n=2). Cells co-expressing multiple markers converge on a single circled cluster (Leiden cluster 22), identified as the ciliated cluster. (F) Isolation of Leiden cluster 22 (resolution=1.5) on the UMAP, comprising 61 cells (0.27% of total PBMCs), highlighted in red against the grey background of all other cells. (G) Dot plot of the 13-gene ciliated module score across all annotated cell types. Dot size represents the percentage of cells with a positive score; colour indicates the mean module score. The ciliated cluster (Leiden 22) shows markedly higher score and expression prevalence than all immune populations. (H) Spatial distribution of the 13-gene ciliated module score projected onto the UMAP. High scores are restricted to the discrete ciliated cluster, with negligible signal across immune populations. (I) UMAP with re-annotated ciliated cells (n=61, 0.27% of PBMCs) labelled in red among all other cell types, confirming their spatial isolation from immune clusters. (J) Violin plots of the ciliated module score across all cell types following re-annotation. The ciliated cluster exhibits a substantially elevated score (median > 0) compared to all immune cell types, which score near zero, confirming the specificity of the ciliated transcriptional signature to this population.

**Supplementary Figure 5.**
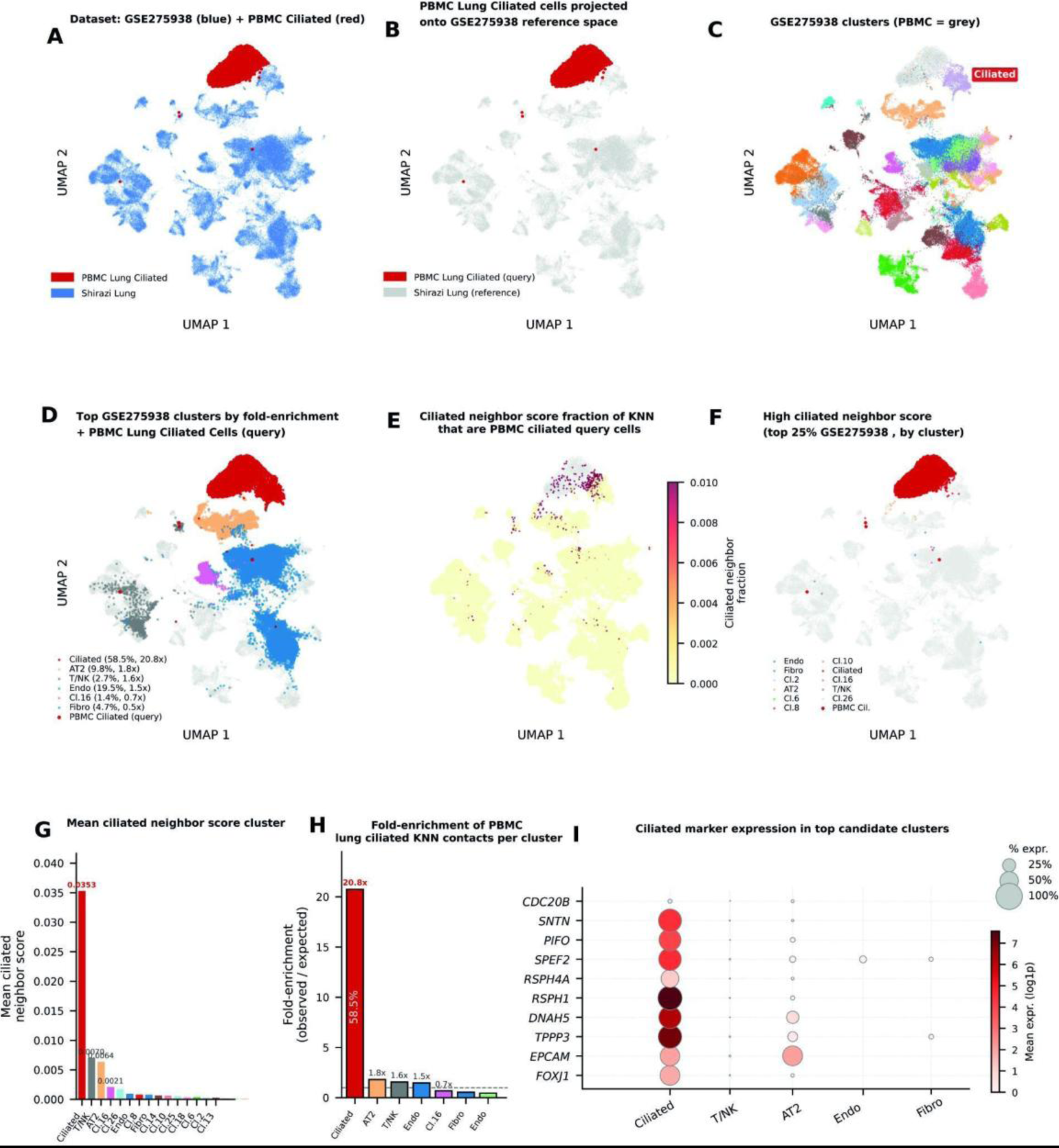
Reference-based integration of PBMC-derived circulating ciliated cells onto the Shirazi et al. neonatal lung atlas (GSE275938) using scArches query mapping. An scVI model was trained on 67,419 neonatal lung cells from GSE275938 (Shirazi et al., 2025) as a reference, and PBMC-derived lung ciliated cells were projected as a query into the established latent space. K-nearest neighbour (KNN) analysis (k=15) was performed to quantify transcriptional proximity between the query cells and each Shirazi cluster. (A) Joint UMAP of the GSE275938 neonatal lung reference (blue, 67,419 cells) and PBMC-derived lung ciliated cells (red) following scArches integration. PBMC ciliated cells form a discrete cluster co-localising with a specific region of the neonatal lung atlas. (B) UMAP of the GSE275938 reference space (grey) with PBMC lung ciliated query cells (red) overlaid, illustrating the spatial projection of circulating cells onto the pre-trained neonatal lung embedding. (C) UMAP of GSE275938 Leiden clusters (PBMC cells shown in grey). The ciliated epithelial cluster is annotated (red box). Colours represent individual Leiden clusters. (D) UMAP highlighting the top GSE275938 clusters ranked by fold-enrichment of KNN contacts with PBMC ciliated query cells (red dots). Legend reports each cluster’s raw percentage of KNN contacts and fold-enrichment over expected frequency based on cluster size. Only clusters above the expected baseline (1.0×) are highlighted. (E) Spatial distribution of the ciliated neighbour score — defined as the fraction of KNN neighbours that are PBMC ciliated query cells — projected onto the GSE275938 UMAP. High scores (yellow–pink) are concentrated within the neonatal lung ciliated cluster. (F) Shirazi cells within the top 25% of ciliated neighbour scores, coloured by cluster identity, overlaid on the full UMAP. PBMC ciliated query cells are shown separately (PBMC Cil.). The signal is predominantly restricted to the ciliated and a small number of adjacent clusters. (G) Bar chart of mean ciliated neighbour score per GSE275938 Leiden cluster, sorted in descending order. The ciliated cluster (red) exhibits a mean score of 0.0353, more than 13-fold higher than the next-ranked cluster, indicating highly specific transcriptional proximity to the PBMC ciliated query cells. (H) Fold-enrichment of PBMC lung ciliated KNN contacts per GSE275938 cluster (all clusters shown, sorted by fold-enrichment). The dashed line indicates the expected frequency (1×) based on cluster size. The ciliated cluster reaches 20.8-fold enrichment with 58.5% of all KNN contacts; all other major lung cell types remain at or below 1.8×. Raw percentage of KNN contacts is indicated below each bar. (I) Dot plot of 10 ciliated marker genes (*FOXJ1*, *EPCAM*, *TPPP3*, *DNAH5*, *RSPH1*, *RSPH4A*, *SPEF2*, *PIFO*, *SNTN*, *CDC20B*) cross the top candidate GSE275938 clusters. Dot size represents the fraction of cells expressing each gene; colour intensity represents mean log1p-normalised expression. All markers are expressed at high levels and in a high fraction of cells exclusively within the ciliated cluster, with negligible expression in T/NK, AT2, endothelial, and fibroblast clusters.

**Supplementary Figure 6.**
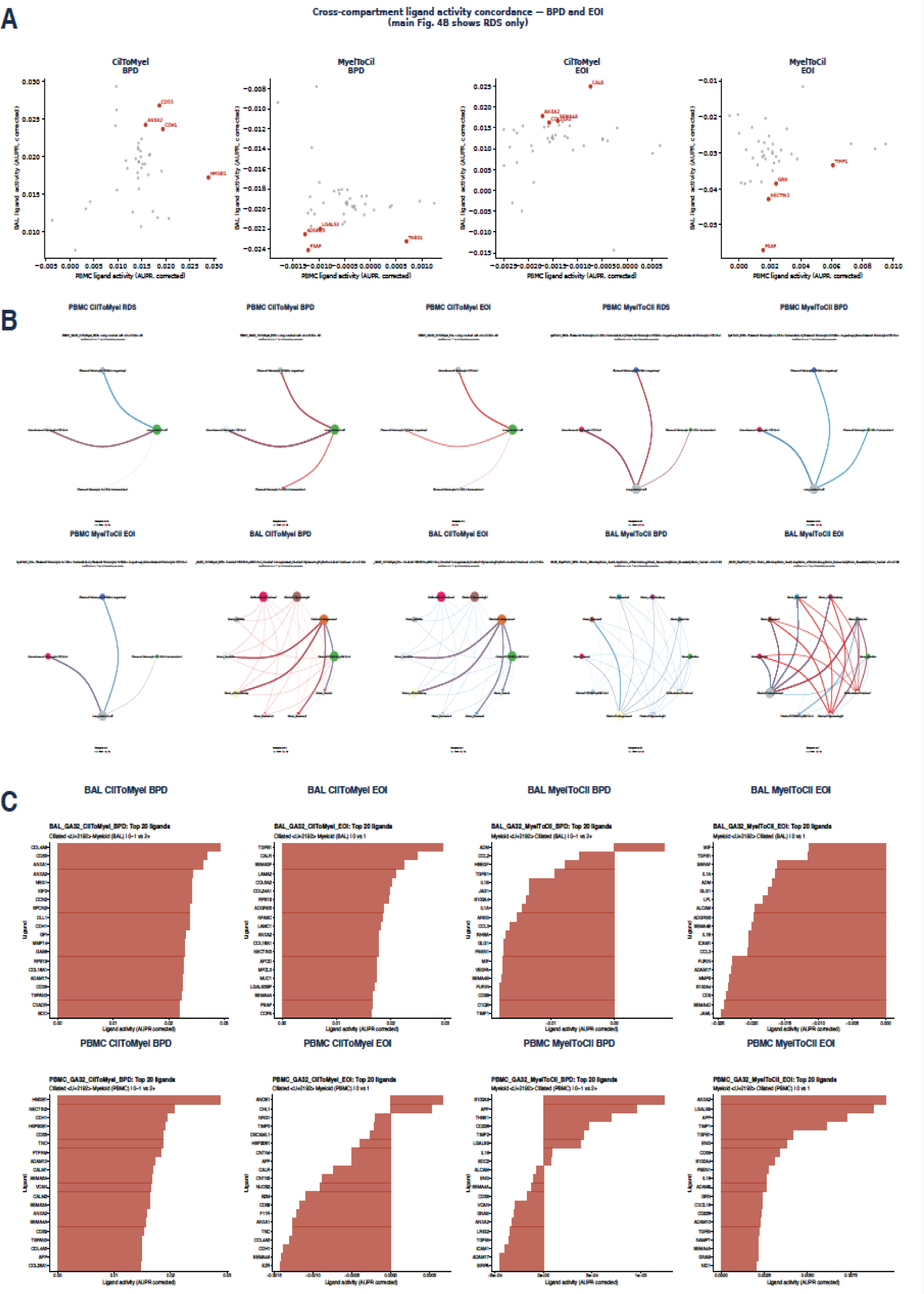
Cross-compartment ligand-receptor signaling concordance between ciliated epithelial and myeloid populations in BPD and EOI. (A) Concordance of ligand activity (AUPR, corrected) between PBMC and BAL/airway compartments for ciliated-to-myeloid (CilToMyel) and myeloid-to-ciliated (MyelToCil) signaling directions, shown for BPD and EOI (RDS shown in the main figure). (B) Circle plots of differential cell-cell communication networks between very and extreme preterm groups in PBMC and BAL compartments, for CilToMyel and MyelToCil directions, in BPD and EOI; edge color denotes receptor expression change (up/down) in extreme preterm infants. (C) Bar plots of the top 20 ligand activities (AUPR, corrected) per compartment (BAL, PBMC), signaling direction (CilToMyel, MyelToCil), and condition (BPD, EOI).

**Supplementary Figure 7.**
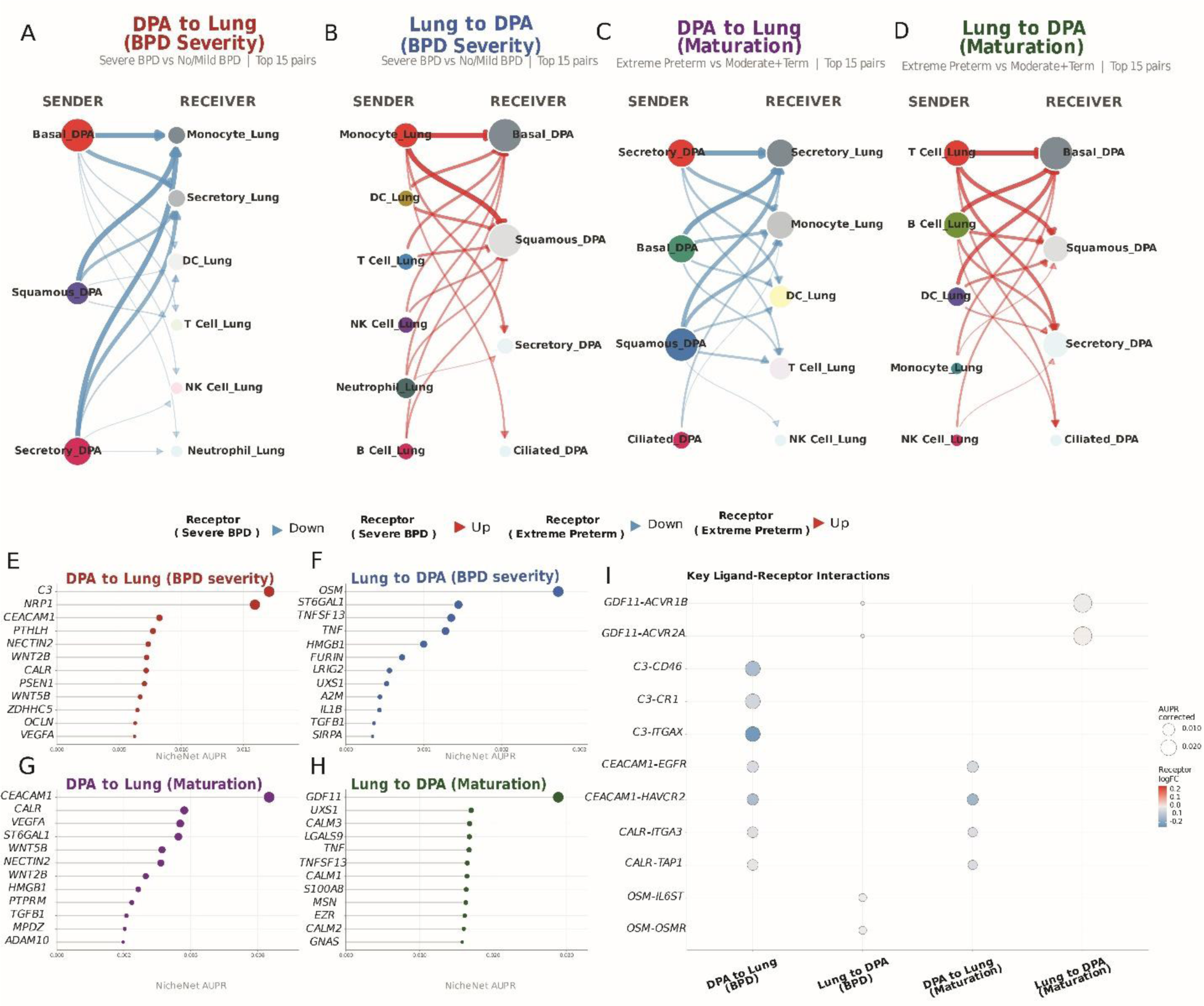
DPA neonatal lung cross-compartment NicheNet analysis predicts disease- and maturation-associated intercellular signaling axes. NicheNet was applied in both DPA to Lung and Lung to DPA directions across four comparisons, using DPA cells from the AIRR cohort and cell types from an independent neonatal lung single-cell atlas (Shirazi et al., GSE275938; 67,419 cells) as the complementary compartment. Analyses were stratified by BPD severity (A1: DPA to Lung; A2: Lung to DPA; Severe BPD vs. No/Mild BPD) and gestational maturity (B1: DPA to Lung; B2: Lung to DPA; Extreme Preterm vs. Moderate and Term). Ligand activity was ranked by NicheNet AUPR corrected score; receptor regulation in the condition of interest was computed as log₂ fold change from pseudobulk differential expression. (A–D) Bipartite sender–receiver communication networks for analyses A1 (A), A2 (B), B1 (C), and B2 (D). Nodes represent cell types, labeled by compartment of origin (_DPA or _Lung); node size is proportional to the number of active sender–receiver interactions. Edge width and opacity reflect summed NicheNet AUPR across all ligand–receptor pairs connecting each sender–receiver combination; edge color indicates the predominant direction of receptor regulation in the condition of interest (red, upregulated; blue, downregulated). The top 15 sender–receiver pairs per analysis are shown. In analysis B2, T Cell_Lung and B Cell_Lung emerge as dominant senders toward DPA epithelial populations, consistent with lymphocyte-to-airway signaling in the maturation context. (E–H) Lollipop plots of the top 12 NicheNet-ranked ligands by AUPR corrected score for each analysis: A1 (E), A2 (F), B1 (G), and B2 (H). Dot size encodes AUPR magnitude. In analysis A1 (DPA to Lung, BPD severity; E), complement component C3 ranked as the top predicted ligand, followed by NRP1, CEACAM1, and CALR. In analysis A2 (Lung to DPA, BPD severity; F), OSM ranked highest by a substantial margin, with TNFSF13 and TNF among the top-ranked signals. In analysis B1 (DPA to Lung, maturation; G), CEACAM1 and CALR were the leading DPA-derived predicted ligands. In analysis B2 (Lung to DPA, maturation; H), GDF11 ranked as the top predicted ligand with the highest AUPR score across all four analyses (AUPR=0.53), consistent with a role for TGF-β superfamily signaling in coordinating lung-to-airway maturation programs. (I) Bubble plot of the top predicted ligand–receptor gene pairs across all four analyses. Each row represents a unique ligand to receptor interaction; each column represents one analysis. Bubble size encodes NicheNet AUPR corrected score; bubble fill encodes mean receptor log₂ fold change in the condition of interest (red, upregulated; blue, downregulated in the disease or preterm group relative to reference; grey, not detected or logFC≈0). GDF11 to ACVR1B and GDF11 to ACVR2A ranked highest in analysis B2, supporting a GDF11–activin receptor axis in lung-to-DPA maturation signaling. C3 to CD46, C3 to CR1, and C3 to ITGAX interactions were predicted in analysis A1, implicating complement-mediated epithelial-to-lung myeloid signaling in BPD severity. CEACAM1 to HAVCR2 and CEACAM1 to EGFR were predicted in analysis B1, extending the immune-checkpoint signaling landscape in extreme prematurity. OSM to IL6ST and OSM to OSMR were predicted in analysis A2, consistent with a pro-inflammatory oncostatin M axis from the neonatal lung toward DPA cells in BPD.

**Supplementary Figure 8.**
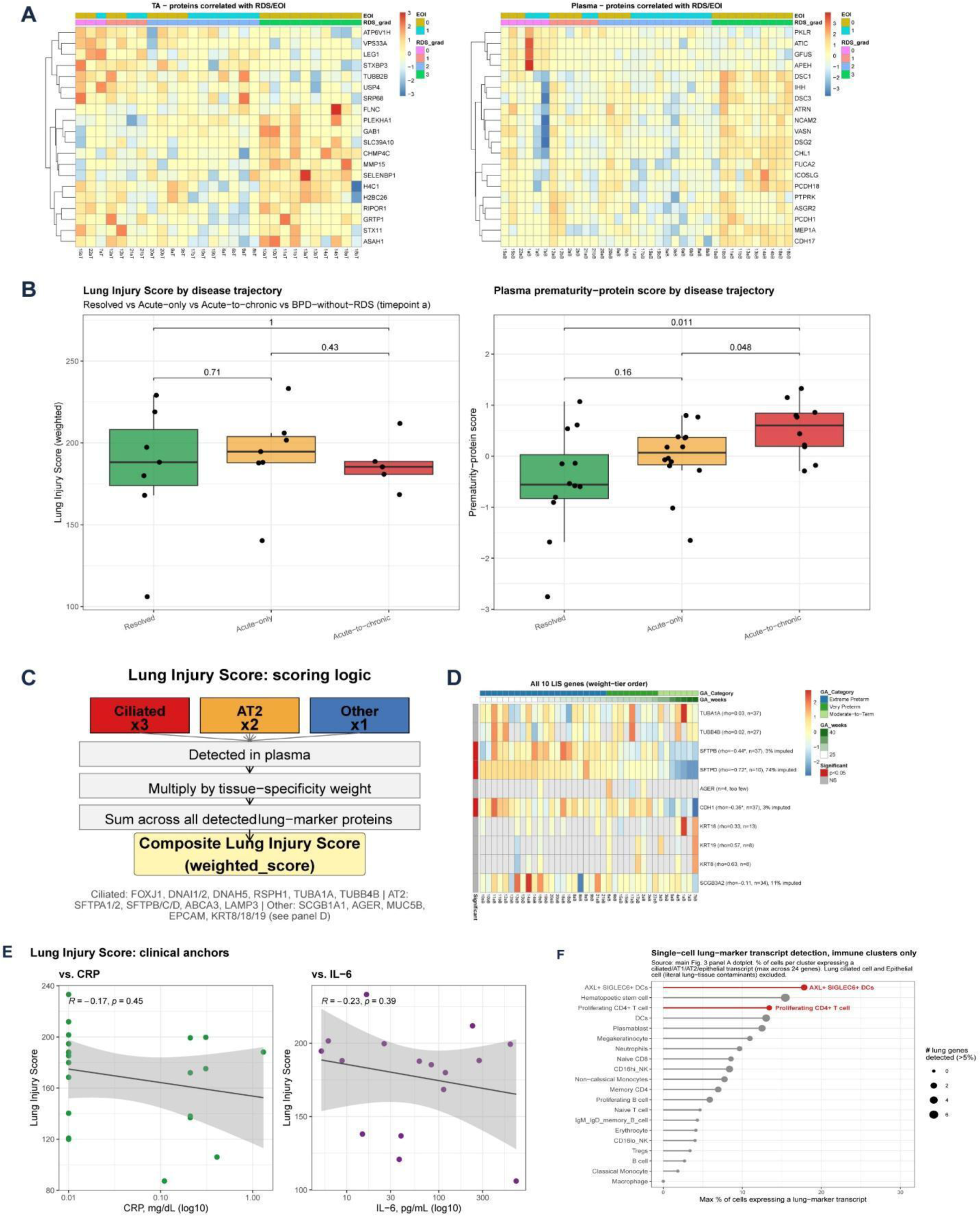
Derivation and validation of the Organ-to-System Score. (A) Heatmaps of tracheal aspirate (DPA, left) and plasma (right) proteins correlated with RDS/EOI severity, annotated by gestational age and BPD/RDS grade. (B) Organ-to-System Score (left) and plasma prematurity-protein score (right) by disease trajectory group (Healthy, Acute-only, Acute-to-chronic, BPD-without-RDS) at Timepoint a; p-values from pairwise comparisons are shown above each bracket. (C) Schematic of the Organ-to-System Score scoring logic: a tissue-specificity-weighted sum (ciliated x3, AT2 x2, other lung markers x1) of detected plasma lung-marker proteins. (D) Heatmap of the 10 lung-marker genes contributing to the score (weight-tier order) across donors and timepoints, annotated by gestational age category and BPD/RDS grade. (E) Correlation of the Organ-to-System Score with clinical inflammatory markers CRP (left) and IL-6 (right); Spearman R and p-value shown. (F) Maximum percentage of cells expressing a lung-marker transcript across single-cell immune clusters, ranked by detection frequency; dot size indicates number of lung genes detected.

**Supplementary Figure 9.**
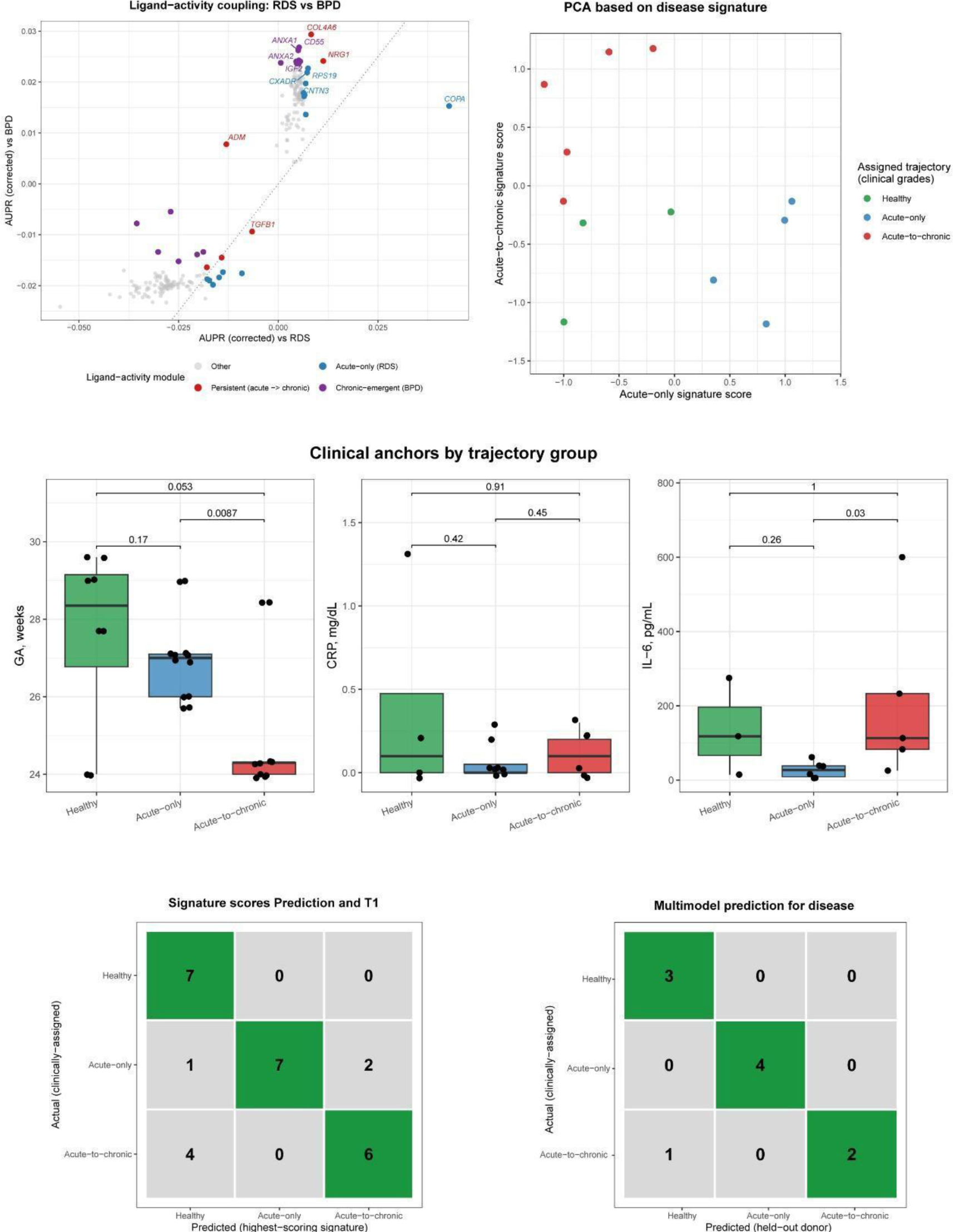
Proteomic disease-trajectory signature scores and outcome prediction model. (A) Concordance of ligand activity (AUPR, corrected) between RDS- and BPD-associated signaling, highlighting persistent (acute→chronic), chronic-emergent (BPD), and acute-only (RDS) ligand-activity modules. (B) Principal component analysis based on the combined disease signature, with donors colored by clinically assigned trajectory (Healthy, Acute-only, Acute-to-chronic). (C-E) Clinical anchors by trajectory group: gestational age at birth (C), CRP (D), and IL-6 (E), compared across Healthy, Acute-only, and Acute-to-chronic groups; pairwise p-values are shown above each bracket. (F) Confusion matrix of disease-trajectory prediction using proteomic signature scores at Timepoint 1, comparing clinically assigned versus highest-scoring predicted trajectory. (G) Confusion matrix of leave-one-donor-out multinomial logistic regression prediction of disease trajectory, comparing clinically assigned versus held-out donor prediction.

